# Oral Baricitinib in the Treatment of Cutaneous Lichen Planus

**DOI:** 10.1101/2024.01.09.24300946

**Authors:** Angelina Hwang, Jacob Kechter, Tran Do, Alysia Hughes, Nan Zhang, Xing Li, Rachael Wasikowski, Caitlin Brumfiel, Meera Patel, Blake Boudreaux, Puneet Bhullar, Shams Nassir, Miranda Yousif, David J DiCaudo, Jennifer Fox, Mehrnaz Gharaee-Kermani, Xianying Xing, Samantha Zunich, Emily Branch, J. Michelle Kahlenberg, Allison C. Billi, Olesya Plazyo, Lam C. Tsoi, Mark R Pittelkow, Johann E. Gudjonsson, Aaron R Mangold

## Abstract

**Background:** Cutaneous lichen planus (LP) is a recalcitrant, difficult-to-treat, inflammatory skin disease characterized by pruritic, flat-topped, violaceous papules on the skin. Baricitinib is an oral Janus kinase (JAK) 1/2 inhibitor that interrupts the signaling pathway of interferon (IFN)-γ, a cytokine implicated in the pathogenesis of LP.

**Methods:** In this phase II trial, twelve patients with cutaneous LP received baricitinib 2 mg daily for 16 weeks, accompanied by in-depth spatial, single-cell, and bulk transcriptomic profiling of pre-and post-treatment samples.

**Results:** An early and sustained clinical response was seen with 83.3% of patients responsive at week 16. Our molecular data identified a unique, oligoclonal IFN-γ, CD8+, CXCL13+ cytotoxic T-cell population in LP skin and demonstrate a rapid decrease in interferon signature within 2 weeks of treatment, most prominent in the basal layer of the epidermis.

**Conclusion:** This study demonstrates the efficacy and molecular mechanisms of JAK inhibition in LP.

**Trial Registration Number**: NCT05188521

## Background

Lichen planus (LP) is a chronic inflammatory condition typified by purple, polygonal, pruritic, papules and plaques.^1^ Cutaneous LP affects 1-2% of the general population and has significant impacts on quality of life (QoL) primarily due to intense pruritus or pain.^2^ Certain subtypes, such as hypertrophic and mucosal LP, are symptomatic, chronic, and refractory to treatment.^3,4^

Treatment of LP is challenging, and therapeutic options have remained largely stagnant. First-line therapy is commonly topical steroids. Other therapies include topical calcineurin inhibitors, oral retinoids, methotrexate, and oral or intralesional steroids.^1^ However, optimal results are rarely achieved, and long-term use of these medications can lead to significant adverse effects. To date, no disease-specific medications have been developed despite the need for therapeutics with a more favorable side-effect profile and for recalcitrant cases.

LP is a T-cell-mediated disease with IFN-γ established as a key mediator.^5^ This cytokine attracts lymphocytes and plasmacytoid dendritic cells to the epidermis and stimulates the interaction between keratinocytes and lymphocytes.^6,7^ CD4+ T-cells release IFN-γ, which leads to CD8+ T-cell stimulation and propagation of the Th1 inflammatory response.^8,9^ Keratinocytes primed by IFN-γ have increased susceptibility to the cytotoxic effects of activated CD8+ T-cells.^8^

IFN-γ signals through the Janus kinase (JAK)-signal transducer and activator of transcription (STAT) pathway, and recent case reports of LP have shown response to JAK inhibitors.^10-15^ An exploratory, open-label study of topical ruxolitinib (JAK-1/2 inhibitor) resulted in significant reductions in total lesion count and modified Composite Assessment Index Lesion Severity (mCAILS) scores, with therapeutic response achieved in 83% of treated lesions.^16^ Baricitinib is an oral JAK-1/2 inhibitor that prevents the phosphorylation of STATs and subsequent signaling of IFN-γ. Case reports and retrospective studies have reported successful treatment of baricitinib in nail LP, oral LP, and lichen planopilaris.^12,17,18^ In this first-in-human trial, we conducted an open-label, single-arm study of baricitinib in cutaneous LP and defined the molecular profile and signature of disease using bulk, spatial, and single-cell RNA-sequencing (scRNA-seq) on pre- and post-treatment specimens.

## Methods

### Trial Design

This single-arm, open-label, phase 2, first-in-human trial was conducted at Mayo Clinic, Arizona (ClinicalTrials.gov identifier NCT05188521). The Mayo Clinic Institutional Review Board approved the study (IRB 21-003075), and all patients provided written informed consent. Twelve patients with biopsy-proven cutaneous LP were administered oral baricitinib 2 mg once daily for 16-weeks. The primary endpoint was an overall response by Physician Global Assessment (PGA) of skin at week 16, with treatment response defined as PGA 0 to 3 (with ≥ 50% score reduction). Secondary outcomes were changes in mCAILS, total body lesion count, affected BSA, pruritus numeric rating scale (NRS), pruritus verbal rating scale (VRS), pruritus visual analog scale (VAS), pain NRS, and Skindex-16. Patients were evaluated at baseline (week 0) and weeks 2, 4, 8, 12, and 16. Treatment-responsive patients who did not achieve PGA grade 0 at week 16 were eligible to enroll in the dose escalation for an additional 12 weeks of treatment with oral baricitinib 4 mg daily. Complete responders at week 16 were reassessed at week 20 and partial responders at week 16 were reassessed at week 32, after an off-therapy period of 4 weeks, respectively.

Assessments of treatment efficacy, adverse events (AEs), and QoL were completed at each study visit. All lesions were annotated, photographed, and scored using the mCAILS criteria.^16^ All clinical assessments were performed by the principal investigator and co-investigators (ARM, MRP, SZ) with questionable lesions scored by 2 investigators. The revised National Cancer Institute Common Terminology Criteria for Adverse Events version 5.0 was used for AE reporting. Due to travel issues, one patient withdrew from the study after week 8. We used intention-to-treat (ITT) analysis with the patient’s last observation to impute week 16 data.

Skin biopsies were collected at baseline and week 2 of LP lesional skin and normal appearing skin for bulk RNA-sequencing. Week 2 samples were designated as responsive (defined as a lesion with ≥ 50% response by mCAILS) or nonresponsive (defined as a lesion with < 50% response by mCAILS).^19^ Additional 6-8 mm biopsies of lesional tissue were taken at weeks 0 and 2 for spatial sequencing and scRNA-seq, and blood for scRNA-seq.

### Eligibility Criteria

Patients aged ≥ 18 years with biopsy-proven cutaneous LP were eligible for the trial. Both treatment-naïve and treatment-refractory disease were included. Key exclusion criteria included predominantly non-cutaneous variants of LP, active infections, and other active inflammatory cutaneous conditions. See supplemental for additional eligibility criteria (Supplementary Note).

### Statistical Analysis

Patient demographics, clinical characteristics, and outcomes were summarized as mean, standard deviation, median, interquartile range for continuous variables, and frequency and percentages for categorical variables. Primary and secondary outcome differences between baseline and week 16 were compared using Wilcoxon signed rank test for continuous variables and McNemar’s test for binary variables. One patient withdrew from the study after week 8, and this patient’s last observation (week 8) was used to impute week 16 data in the ITT analysis. Exact binomial method was used to calculate the treatment response rate at week 16 and its corresponding 95% confidence interval. All analyses were conducted with R version 4.1.2 (R Foundation for Statistical Computing, Vienna, Austria). The cut-off for statistical significance was set as 0.05.

### Transcriptomic Analyses

Bulk RNA sequencing, scRNA sequencing, and spatial sequencing were conducted on pre- and post-treatment timepoints on control and disease samples. T-cell receptor gene expression and cell type annotations were conducted on skin samples to identify clonally expanded T-cell types. Immunohistochemical staining of CD3, IFN-γ, CXCL9, and pSTAT2 were performed on skin samples. Detailed methods for transcriptomic analyses, TCR clonality analyses, and immunohistochemistry are provided in the Supplemental Methods.

## Results

### Patients

A total of 12 patients with a mean age of 63.6 (SD 13.6) years were enrolled. The majority (n=11, 91.7%) were female and identified as White (n=9, 75.0%). Underrepresented minority patients constituted the remaining 25%. The mean disease duration across all patients was 26.5 months (SD 30.8). All patients had treatment-refractory LP, with 91.7% failing topical steroids, 41.7% failing oral and intramuscular steroids or non-steroidal immunosuppressants, 8.3% failing methotrexate, and 8.3% failing topical calcineurin inhibitors. Classic LP was the predominant form, with hypertrophic LP seen in five (41.7%) patients and mucosal involvement in two subjects (16.7%).

The average affected body surface area (BSA) was 4.9% at baseline (SD 3.7) with a mean of 151.9 total body LP lesions per patient (range 4 – 600). The mean baseline mCAILS score was 12.3 (SD 3.2) and the overall Skindex-16 was 59.0 (SD 22.1). Pruritus NRS and pruritus VAS scores were 7.2 (SD 2.4) and 6.6 (SD 1.6), respectively, with 91.7% of patients rating their level of itch as moderate/severe on the pruritus VRS. The baseline pain NRS score was 7.7 (SD 1.7).

### Efficacy

At week 16, 10 of 12 (83.3%; 95% CI: 51.6% - 97.9%) patients demonstrated treatment response, achieving PGA scores of 0 to 3, with ≥ 50% score reduction (Figure 1; Table 1). Five of the 10 treatment-responsive patients had a PGA of 0 (completely clear), and five had a PGA of 1 (almost clear) (Supplementary Table 1; Supplementary Figure 1). Improvement in PGA was observed as early as week 1 in 37.5% of patients and in 100% of patients by week 12. Treatment effects were sustained at week 20 (4 weeks off-therapy), with all patients demonstrating continued response.

**Figure 1:**
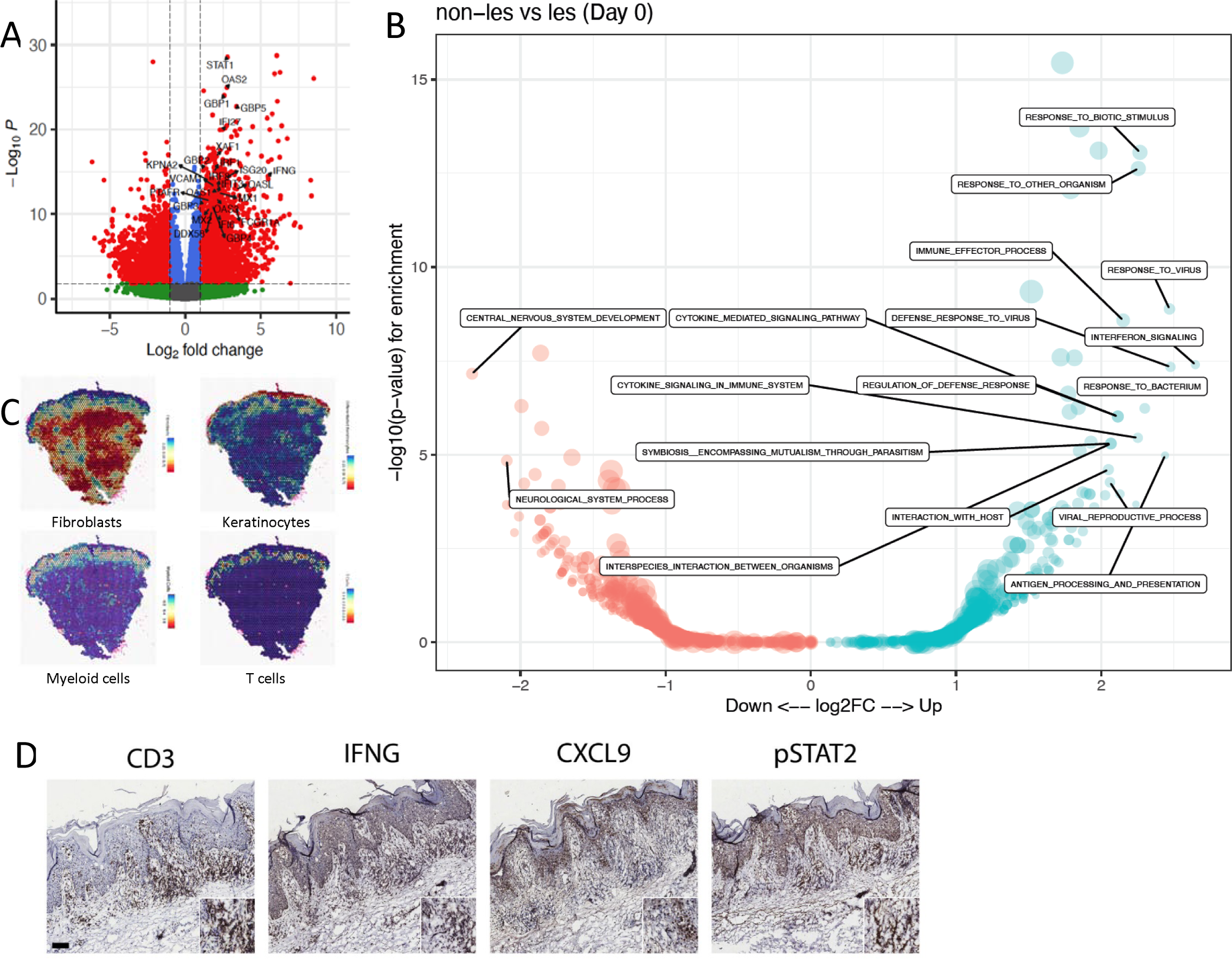
LP is an IFN driven disease process. (A) Volcano plot of bulkRNA-seq data comparing lesional vs. nonlesional LP skin at Day 0 (n=10 and 9, respectively) (red color shows FDR<0.05 and Log2 FC>1 or <-1, blue is FC<1 and >-1 and FDR<0.05), green is FC>1 or <-1 and FDR>0.05). (B) Enriched GO categories in DEGs between lesional vs. nonlesional LP skin (red, and blue colors represent enriched GO categories amongst increased vs. decreased DEGs, respectively, p<0.05). (C). Cellular deconvolution of fibroblasts, keratinocytes, myeloid cells, and T cells on the Visium 10X spatial expression platform (representative of n=9). (D) IHC of the T cell marker CD3, pSTAT2, IFN-γ, and CXCL9 (representative of n=9, scale bar = 100um).

**Table 1.** Primary and secondary endpoints at baseline and week 16 (intention-to-treat analysis)

|  | **Baseline** | **Week 16** | **Difference** | **P value** |
| --- | --- | --- | --- | --- |
| PGA |  |  |  |  |
| Responsive (n, %) |  | 83.3% |  |  |
| Non-responsive (n, %) |  | 16.7% |  |  |
| Total Body Lesion Count |  |  |  | 0.002 |
| Mean (SD) | 151.9 (162.1) | 17.1 (33.5) | -134.8 (157.0) |  |
| Range | 4.0 – 600.0 | 0.0 – 117.0 | -570.0 to -4.0 |  |
| mCAILS |  |  |  | 0.002 |
| Mean (SD) | 12.3 (3.2) | 1.7 (3.3) | -10.6 (2.9) |  |
| Range | 7.0 – 18.6 | 0.0 – 8.9 | -14.6 to -5.0 |  |
| BSA affected (%) |  |  |  | 0.002 |
| Mean (SD) | 4.9 (3.7) | 1.0 (2.5) | -3.9 (2.9) |  |
| Range | 0.1 – 12.0 | 0.0 – 8.5 | -9.8 to -0.1 |  |
| Pruritus NRS |  |  |  | 0.003 |
| Mean (SD) | 7.2 (2.4) | 1.8 (3.2) | -5.3 (2.6) |  |
| Range | 1.0 – 10.0 | 0.0 – 10.0 | -8.0 to -0.0 |  |
| Pruritus VAS |  |  |  | 0.003 |
| Mean (SD) | 6.6 (1.6) | 1.7 (3.0) | -5.0 (2.6) |  |
| Range | 2.7 – 8.8 | 0.0 – 9.0 | -7.7 to 1.5 |  |
| Pain NRS |  |  |  | 0.005 |
| Mean (SD) | 7.7 (1.7) | 1.9 (3.2) | -5.6 (2.7) |  |
| Range | 4.0 – 10.0 | 0.0 – 10.0 | -8.0 to 0.0 |  |
| Skindex-16 overall |  |  |  | 0.008 |
| Mean (SD) | 59.0 (22.1) | 16.9 (28.7) | -37.3 (18.3) |  |
| Range | 35.0 – 96.0 | 0.0 – 96.0 | -58.0 to 0.0 |  |
| Skindex-16 symptom |  |  |  | 0.005 |
| Mean (SD) | 16.4 (6.0) | 3.7 (7.0) | -12.4 (5.8) |  |
| Range | 4.0 – 24.0 | 0.0 – 24.0 | -19.0 to 0.0 |  |
| Skindex-16 emotional |  |  |  | 0.003 |
| Mean (SD) | 29.9 (9.0) | 9.0 (12.4) | -20.9 (9.4) |  |
| Range | 17.0 – 42.0 | 0.0 – 42.0 | -34.0 to 0.0 |  |
| Skindex-16 functional |  |  |  | 0.012 |
| Mean (SD) | 11.6 (10.2) | 4.5 (9.0) | -5.5 (5.6) |  |
| Range | 0.0 – 30.0 | 0.0 – 30.0 | -15.0 to 0.0 |  |

Improvements were seen across all secondary measures at week 16 (Table 1). The mean total body lesion count decreased to 17.1 (SD 33.5; p=0.002), and the mean affected BSA decreased to 1.0 (SD 2.5; p=0.002). Compared to a baseline score of 7.2 (SD 2.4), pruritus NRS decreased to 1.8 (SD 3.2; p=0.003) (Supplementary Figure 2), pruritus VAS decreased from 6.6 (SD 1.6) to 1.7 (SD 3.0; p=0.003), and pain NRS decreased from 7.7 (SD 1.7) to 1.9 (SD 3.2; p=0.005) (Supplementary Figure 3). Pruritus NRS improvement ≥ 4 from baseline (NRS4) and Pain NRS4 were achieved in 75.0% and 66.7% of patients, respectively. The overall Skindex-16 score decreased from baseline to week 16 by a mean of 37.3 (SD 18.3; p=0.008), accompanied by decreases in each Skindex subscore: Symptom -12.4 (SD 5.8; p=0.005), Emotional -20.9 (SD 9.4; p=0.003), Functional -5.5 (SD 5.6; p=0.012). Results from the per-protocol analysis, with the population defined as patients who completed 16 weeks of baricitinib, were consistent with the results of the ITT analysis (Supplementary Table 2).

### Dose Escalation

Five of six eligible patients participated in the dose escalation period. All five patients completed an additional 12 weeks of treatment with baricitinib 4 mg daily. At the primary endpoint of week 16, corresponding with the start of dose escalation, 80.0% of patients had PGA grade 1, and 20% had PGA grade 4. After 12 weeks of baricitinib 4 mg daily, 60.0% of patients were completely clear of disease (PGA grade 0), 20.0% were almost clear (PGA grade 1), and 20.0% had slight improvement (PGA grade 4) (Supplementary Table 3). All but one patient remained treatment-responsive upon re-evaluation after 4 weeks off therapy (Supplementary Table 4).

### Safety

There were a total of 12 AEs, with only one mild AE that was deemed probably related to the study drug (absolute neutrophil count 0.78 x 10(9)/L) (Supplementary Table 5). Most AEs were mild or moderate (58.3% and 25.0%, respectively). No AEs led to the discontinuation of baricitinib.

### Molecular Profiling

Whole transcriptomic bulk RNA-seq was performed on lesional and non-lesional skin prior to therapy (n=11, 12, respectively). Analysis for differentially expressed genes (DEGs) (False Discovery Rate, FDR ≤ 0.05, and Fold Change (FC) ≥ 2), revealed a total of 3,524 DEGs, with 1,683 increased and 1,841 decreased compared to non-lesional LP skin at baseline. The most prominent DEGs in lesional LP skin were interferon-stimulated genes (ISGs), including *STAT1* (FC=6.9, FDR=8×10^-25^), *OAS2* (FC=6.7, FDR=9×10^-22^), *MX1*(FC=4.8, FDR=1×10^-10^), and *ISG20* (FC=6.6, FDR=2.9×10^-12^), with *IFN-*γ (FC=44.6, FDR=2.0×10^-12^) (Figure 1A). No differences were observed in the expression of type I IFN genes. Enriched biological processes included, *immune-effector process*, *response to virus*, *interferon signaling*, and *antigen processing and presentation* (Figure 1B). Spatial sequencing data from lesional LP showed dense myeloid and T-cells signatures in the upper dermis adjacent to the epidermis (Figure 1C), and validated by immunohistochemistry (Figure 1D).

Early tissue transcriptomic response was seen after only two weeks of baricitinib treatment, with marked suppression of interferon stimulated genes (Figure 2A), and interferon signaling (p=1.2×10^-15^). This was supported by single-cell analyses comparing early response to baricitinib (2 weeks) to baseline disease. Thus, shifts were observed in the myeloid and T-cell clusters (Figure 2B, 2C, Supplementary Figure 4A and 4B). Keratinocytes, particularly in the basal layer of the epidermis, are the cellular target of T-cell responses in LP skin. ^8^ Basal layer keratinocytes (*KRT5*+) demonstrated two distinct states, “basal KC 1” and “basal KC 2” (Figure 2D, Supplementary Figure 5A-D), with the “basal KC 1” state having an inflammatory state enriched for interferon and JAK1/JAK2 signaling and expression of MHC class I and class II molecules (Figure 2F), which were absent in the “basal KC 2” state (Figure 2G, Supplementary Figure 5C), suggesting that basal KC 1 keratinocytes are the main target of cytotoxic responses in LP. Strikingly, within only 2 weeks of baricitinib treatment there was a marked shift from “basal KC 1” state to “basal KC 2” state (Figure 2E), likely reflecting suppression of IFN responses (Figure 2F, Supplementary Figure 5D) and decreased antigen presentation (Figure 23G).

**Figure 2:**
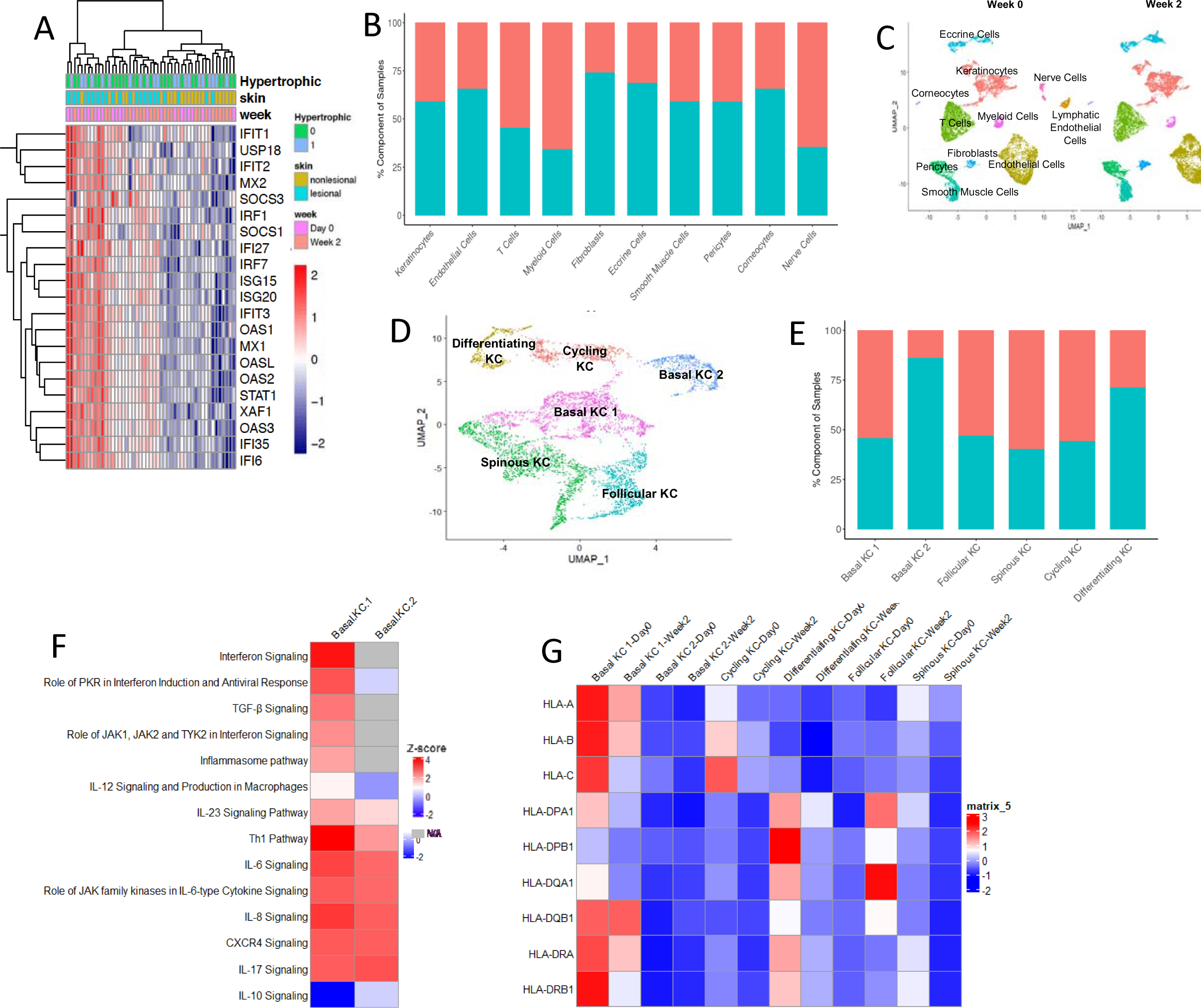
Cellular composition of LP and effect of baricitinib treatment. (A) Changes in gene expression in interferon signature genes at baseline and week 2. (B) Cell proportions at baseline and week 2. (C) Single-cell data from baseline (Day 0) and at week 2 in the patient cohort (n=10, 10). (D) Single-cell data from the LP cohort defines 6 distinct keratinocyte clusters, including two basal cell states. (E) The proportion of each keratinocyte subset at baseline and week 2 of treatment. (F) Enriched GO categories in the two basal keratinocyte clusters. (G) expression of MHC class I and class II molecules in the different keratinocyte compartments at different time points.

Prominent T-cell infiltration is a hallmark feature of LP histology, but the nature of the T-cell involvement has not previously been addressed. We identified six subsets of T-cells in LP skin, including T regulator cells (T regs), CD4+ central memory T-cells, “stressed” T-cells, CD8 cytotoxic T-cells, gamma-delta T-cells, and a novel *CXCL13*+ T-cell subset (Figure 3A, 3B, Supplementary Figure 6A). CD8 cytotoxic, gamma-delta, and *CXCL13*+ T-cell subsets were the main source of *IFN-*γ expression in LP skin (Figure 3C), and demonstrated prominent cell-cell interactions with stromal cells, particularly basal keratinocytes (Figure 3D). The T-cell subsets were localized at the dermal-epidermal junction with prominent expression of cytotoxic markers including *GZMB*, *GZMA*, and *GNLY* (Supplementary Figure 6B). Consistent with the T-cell response being directed against self-antigen(s), the *CXCL13+* CD8+ subset had evidence of oligoclonality in LP skin (Figure 3E), and we confirmed the proximity of CXCL13 T cells to the epidermis in inflamed LP epidermis (Figure 3F). We did not observe prominent mRNA expression of other T-cell cytokines in LP, including the Th17 cytokines *IL17A*, *IL17F*, *IL22*, and *IL26* or the Th2 cytokine *IL4* (Supplementary Figure 7). Interestingly, we observed IL13 mRNA expression in central memory CD4+ T cells and the CXCL13 T cell subset, and in our bulk RNA-seq data (17.6-fold, FDR=3.3×10^-08^).

**Figure 3:**
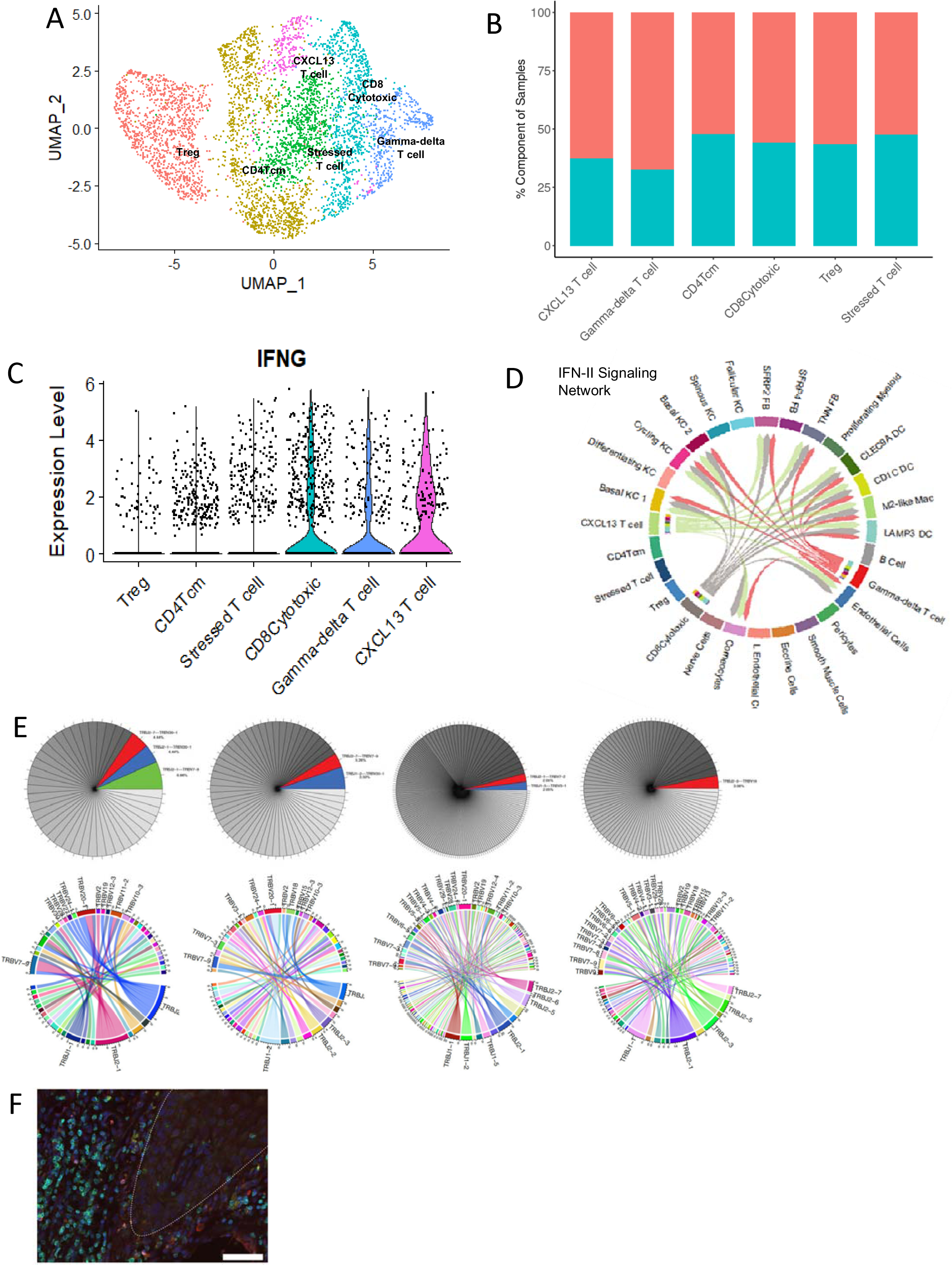
T cell function in LP. (A) Six T cell subsets are found in LP skin. (B) The proportion of T cell subsets at baseline and week 2 of treatment. (C) IFNG expression in T cells subsets in LP skin. (D) Type II IFN signaling network in LP skin. (E) Oligoclonality of CXCL13+ CD8+ T cells in LP skin (3 representative patients). (F) Immunofluorescence of CD3 (green) and CXCL13 (red) in LP skin, showing co-localization of double-positive CXCL13+-CD8+ T cells adjacent to the epidermal-dermal junction (white broken line) (representative image of n=3).

Five myeloid cell populations were identified in LP skin (M2-like, *LAMP3*, *CD1C*, *CLEC9A*, and proliferating myeloid cells) along with a small number of B-cells (Supplementary Figure 8A) and three major fibroblast subsets (*SFRP2*, *TNN*, and *SFRP4*) (Supplementary Figure 8B).

Consistent with LP being a systemic disease, we observed a decrease in ISG expression, including *IFITM1*, MHC class I (*HLA-B*), class II (*HLA-DPA1*), and the cytotoxic marker *GNLY*, with baricitinib treatment across multiple cell populations (Supplementary Figure 9A), along with decrease in biological processes including *interferon signaling* (p=1.5×10^-8^) in myeloid cells and *MHC class II antigen presentation* (p=9.4×10^-4^) in CD4+ T-cells (Supplementary Figure 9C), but no major shifts in cell populations was observed (Supplementary Figure 9B).

## Discussion

This open-label, single-arm trial demonstrated rapid and sustained response to baricitinib in cutaneous LP. Notably, the majority of the patients in our study had chronic, treatment-refractory LP, with half having failed systemic therapy, including methotrexate and oral or intramuscular corticosteroids. This rapid response included improvements in all measures of disease activity including physician global assessment, quality of life measures and pruritus^20,21,22^. Only five patients required dose escalation of baricitinib to 4mg daily. Although evaluation with a larger cohort is needed to establish the safety and efficacy of this increased dosing, our data suggest a dose-dependent response to baricitinib in LP.

The underlying mechanism of pruritus in LP remains unknown, but similar responses were seen in a trial with topical ruxolitinib, a JAK1/2 inhibitor^16^ Th2 cytokines (IL-4, IL-13, and IL-31) are established as key mediators in chronic pruritus. ^23,24^ Among these, IL-4 has been demonstrated to enhance responsiveness to pruritogens via JAK1 phosphorylation. ^25^. Strikingly, while the pathologic responses in LP are heavily skewed towards Th1 responses, IL-13, but not IL-4, was found to be elevated in LP skin, suggesting that it’s immunopathogenesis is more complex than a simple Th1 polarization response, and may suggest that IL-13 may have a pro-pruritic role in LP, which is effectively ameliorated by baricitinib.

Our data suggests that LP is an autoimmune disease with T cell responses directed against a pathogenic self-antigen. It has been suggested that T-cells are central disease mediators, with cytotoxicity mediated through IFN-γ priming of keratinocytes through MHC class I induction and cell death.^8^ Consistent with those findings, the primary source of IFN-γ in LP is cytotoxic CD8+ and gamma-delta T-cells. Here, we identify a novel subset of *CXCL13*+ CD8+ T-cells that are a major source of IFN-γ in LP and show oligoclonality, suggesting reactivity against a limited set of possible autoantigens. Similar *CXCL13*+ CD8+ T-cells have been described for tumor-reactive cells triggered by immune-checkpoint blockade. ^26^ Interestingly, lichenoid dermatitis is not an uncommon cutaneous side-effect of immune-checkpoint inhibitor treatment ^27^, although the specific T-cell lymphocyte population in that setting has not been previously explored. Notably, the three CD8+ T-cell populations were localized at the dermal-epidermal junction and showed predicted interactions with basal layer keratinocytes where enriched IFN responses, and MHC class I and class II expression were observed. These findings are consistent with IFN-γ signaling priming basal cells towards cytotoxic attack, setting the stage for a vicious self-sustaining cycle of cytotoxic responses against basal keratinocytes. Literature-based network analysis of genes demonstrated this signaling to be dependent on JAK/STAT signaling. ^8^ Consistent with this scenario, MHC class I expression rapidly decreases in basal keratinocytes with baricitinib treatment suggesting that baricitinib acts by protecting keratinocytes from IFN-γ induced cytotoxic responses.

We did not observe evidence of involvement of IL-17 in LP pathogenesis, as previously suggested. ^28^ However, most of the reports on IL-17 in LP have focused on oral LP, which was not included in our clinical trial.

Serious adverse events did not occur with baricitinib. A single AE of neutropenia was deemed probably related to the study drug, yet this was mild and did not result in treatment discontinuation. Based on the promising results of this open-label, single-arm trial, future randomized controlled trials of baricitinib are warranted.

## Data Availability

Data produced in the present study are available upon reasonable request to the corresponding author

## Acknowledgements

Aubrey Smith

