## Supplementary Figure 1 for "Oral Baricitinib in the Treatment of Cutaneous Lichen Planus"

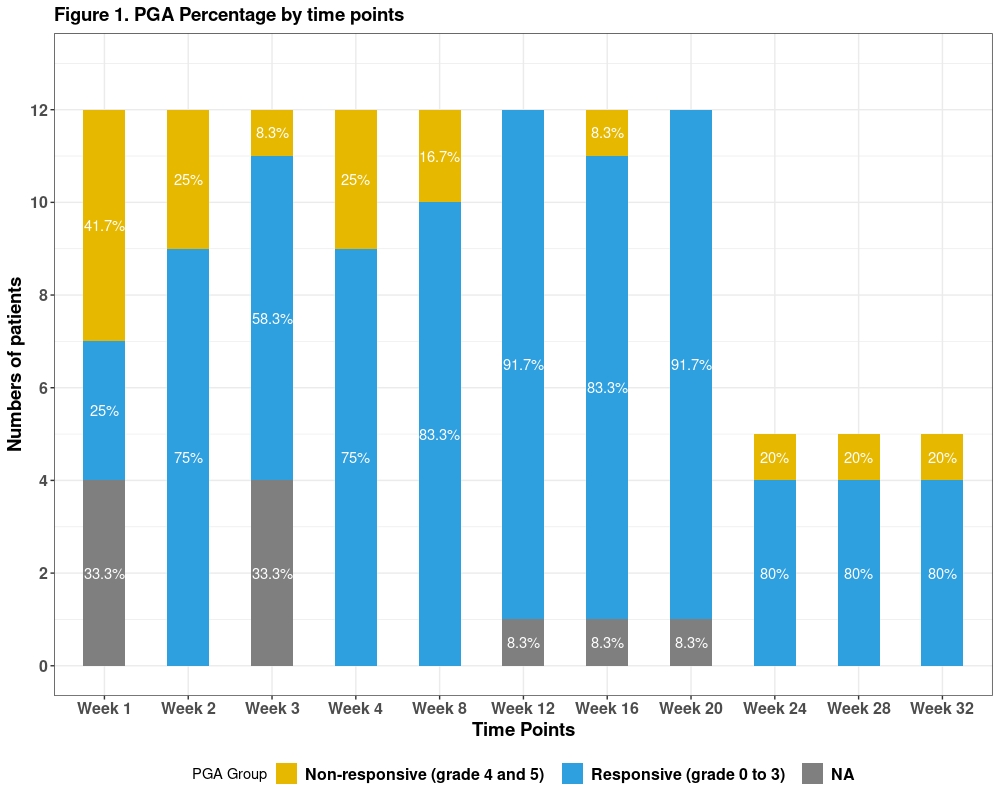

**Supplementary Figure 1**. Treatment response rates as defined by Physician Global Assessment (PGA) from baseline to week 16.
