## Supplementary figures and images for "Oral Baricitinib in the Treatment of Cutaneous Lichen Planus"

### Supplementary Figure 2

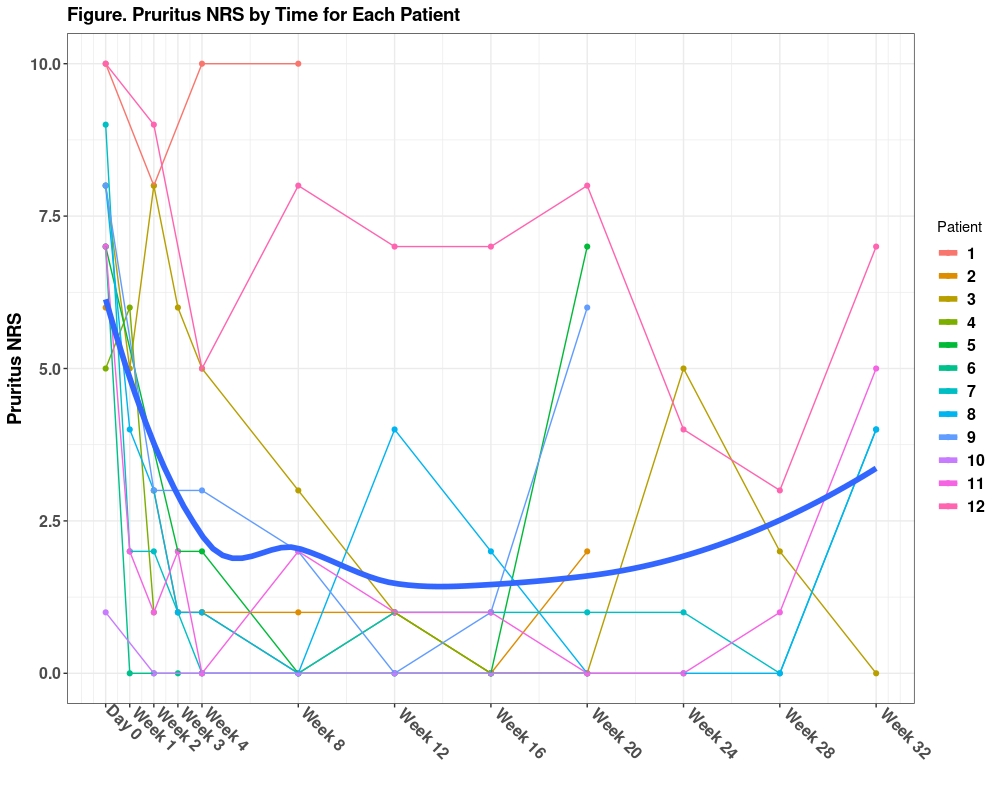

**Supplementary Figure** **2**. Pruritus NRS by time for each participant from baseline to week 32.

### Supplementary Figure 3

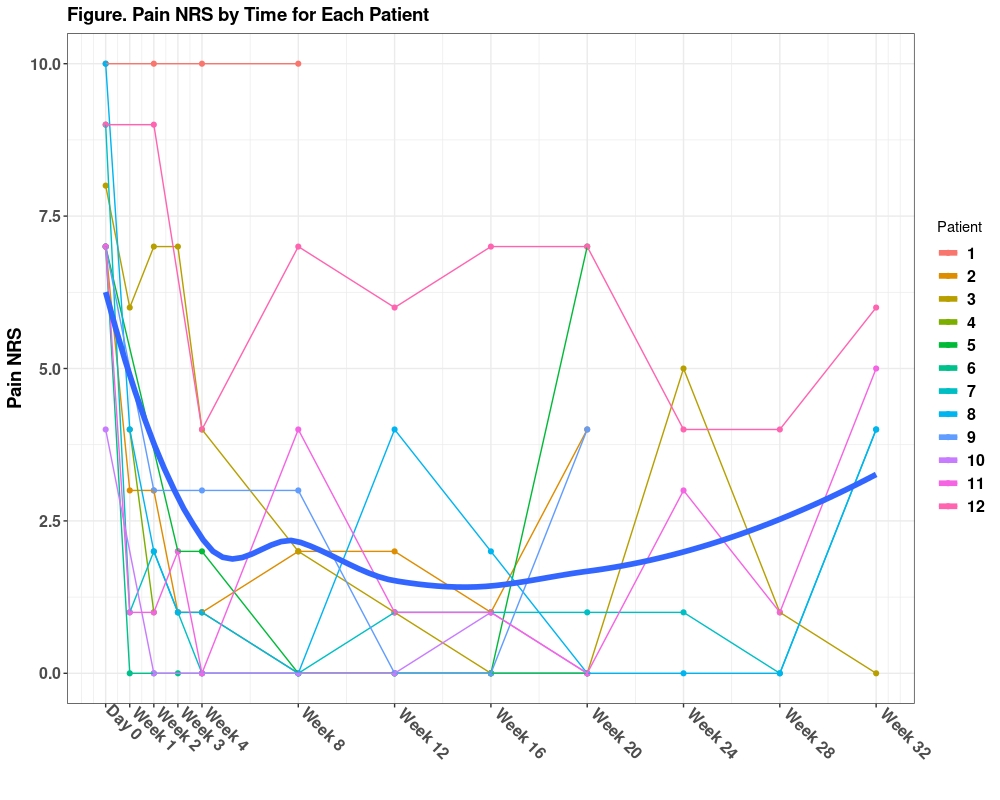

**Supplementary Figure 3.** Pain NRS by time for each participant from baseline to week 32.

### Supplementary Figure 7

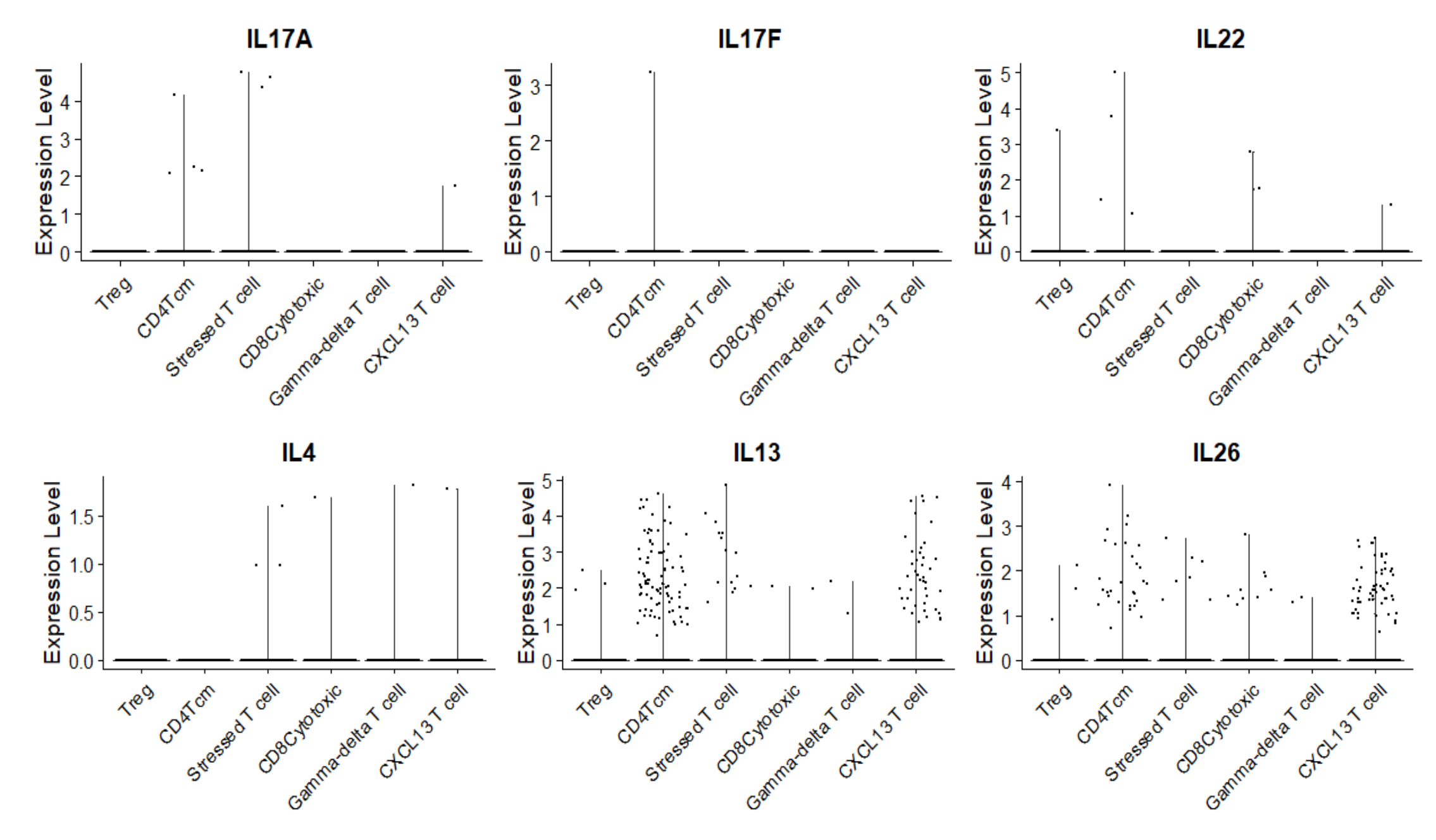

**Supplementary Figure 7.** Expanded panel of cytokine expression in lesional LP T cells.
