## Supplementary Figure 4 for "Oral Baricitinib in the Treatment of Cutaneous Lichen Planus"

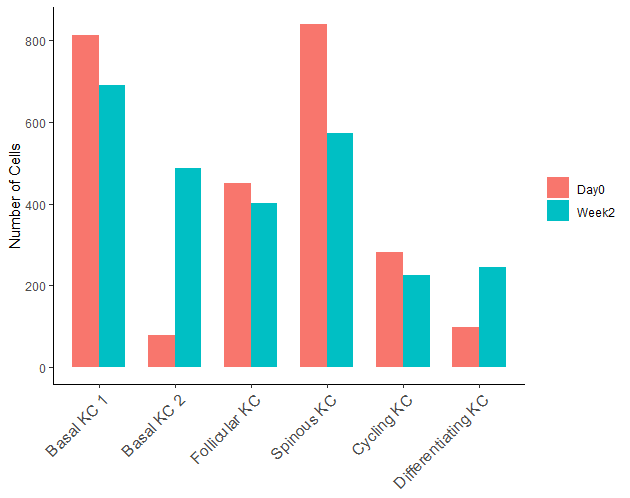

Week 0

Week 2

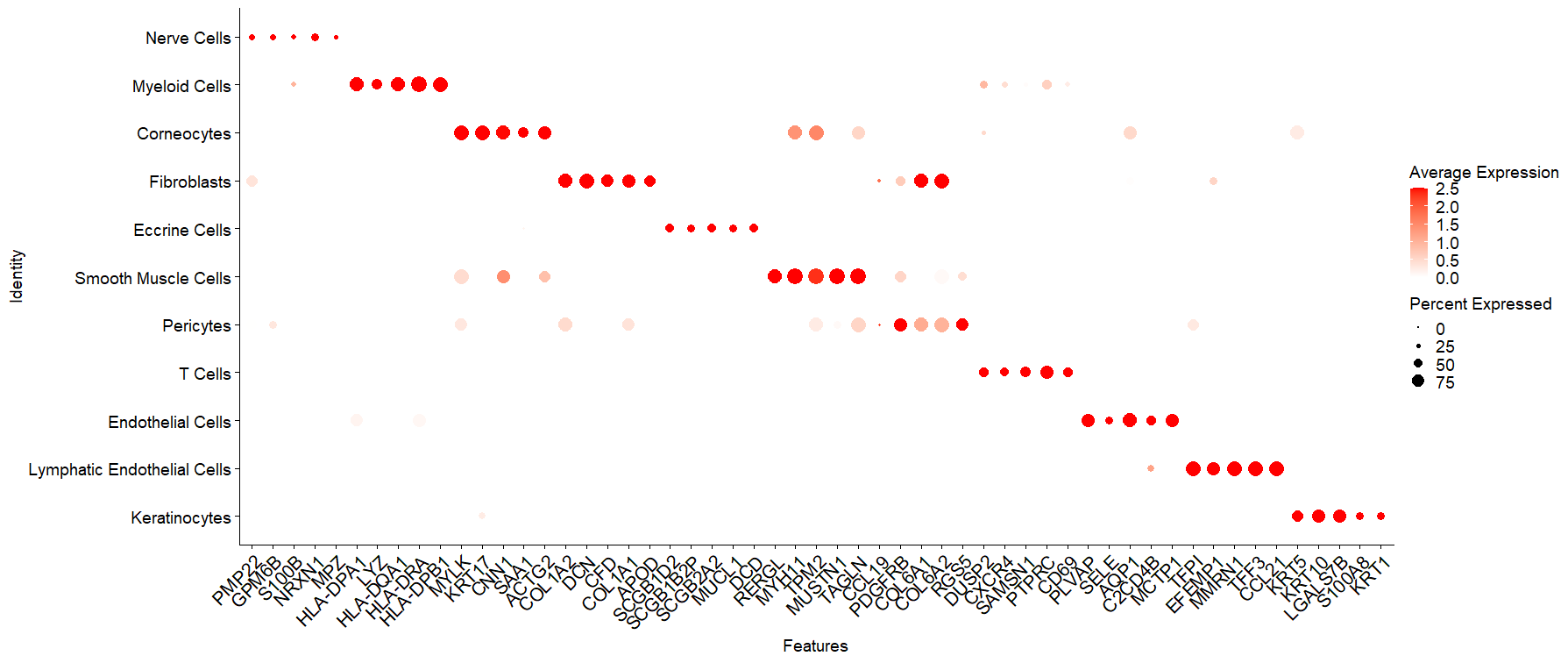

A

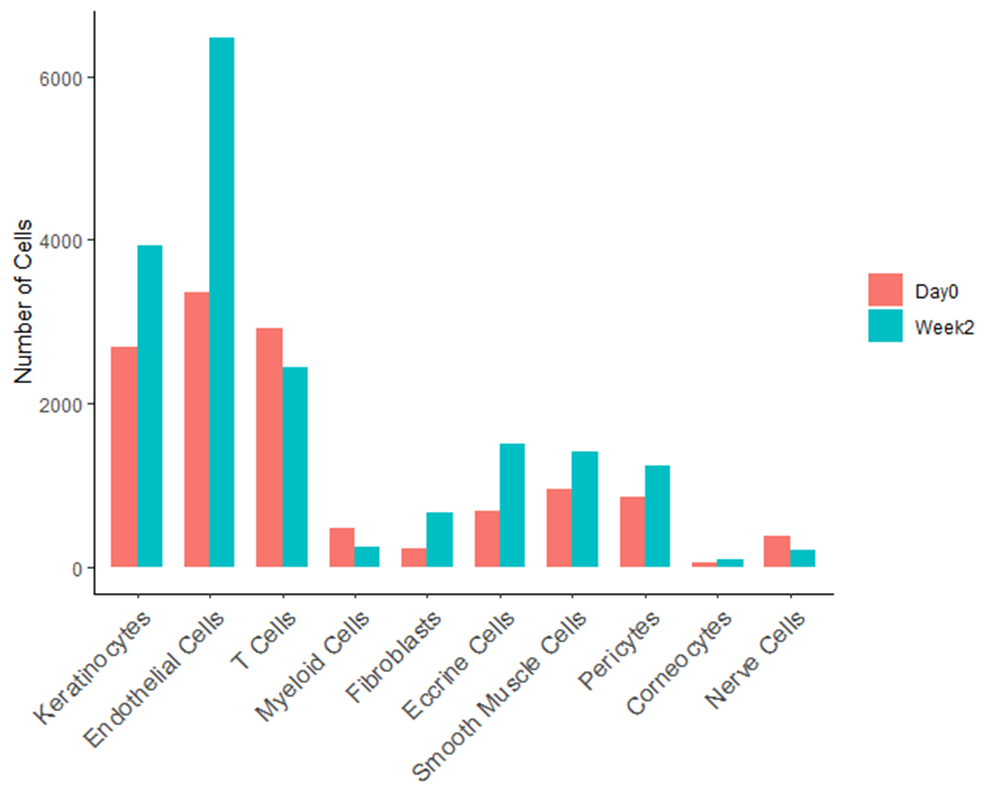

B

**Supplementary Figure 4.** (A) Top 5 gene markers in major cell types in lesional LP skin. (B) Cell numbers at baseline (week 0, red) and week 2 (blue) (n=10, 10).
