## Supplementary Figure 5 for "Oral Baricitinib in the Treatment of Cutaneous Lichen Planus"

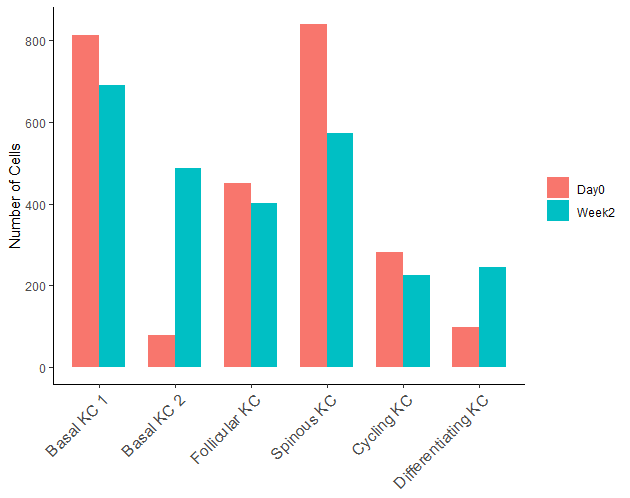

Week 0

Week 2

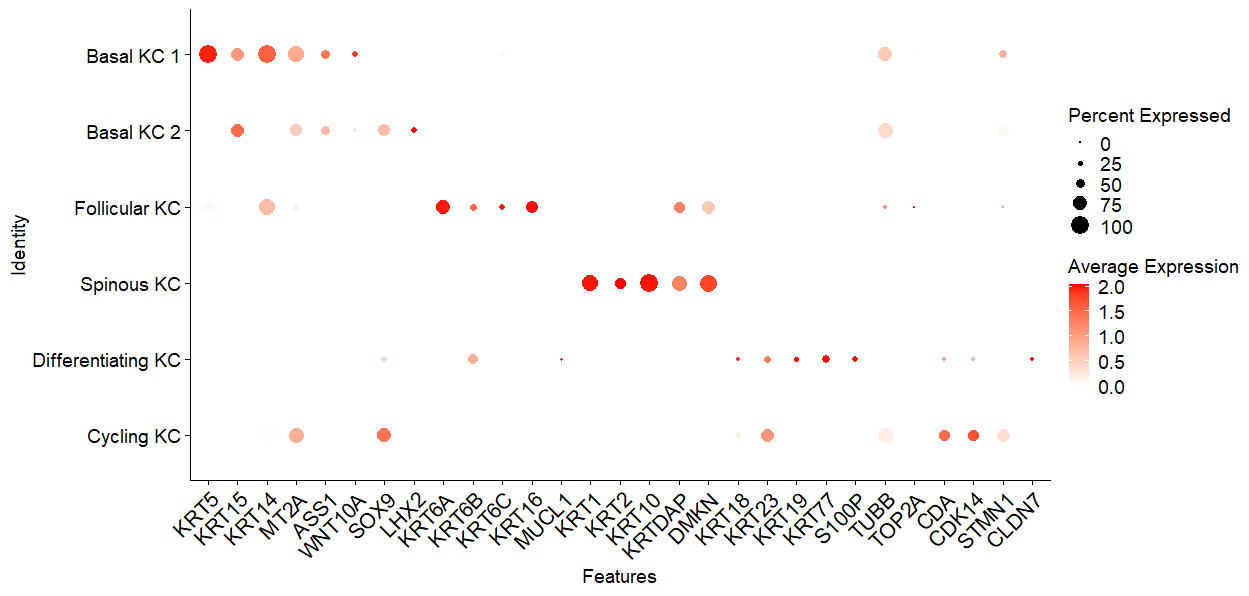

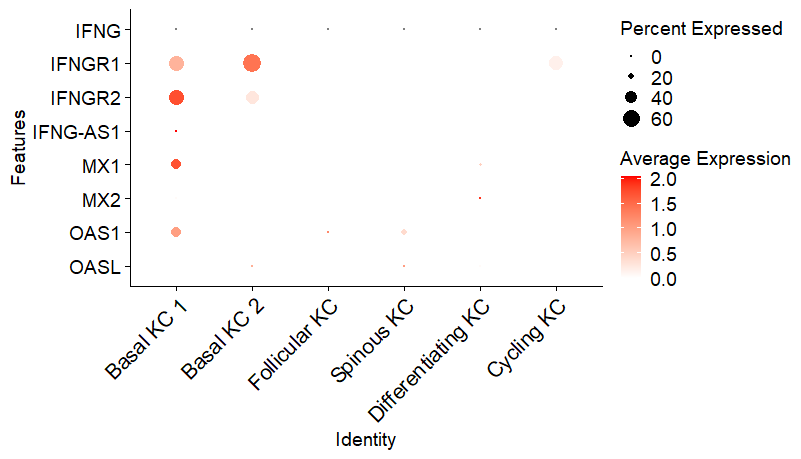

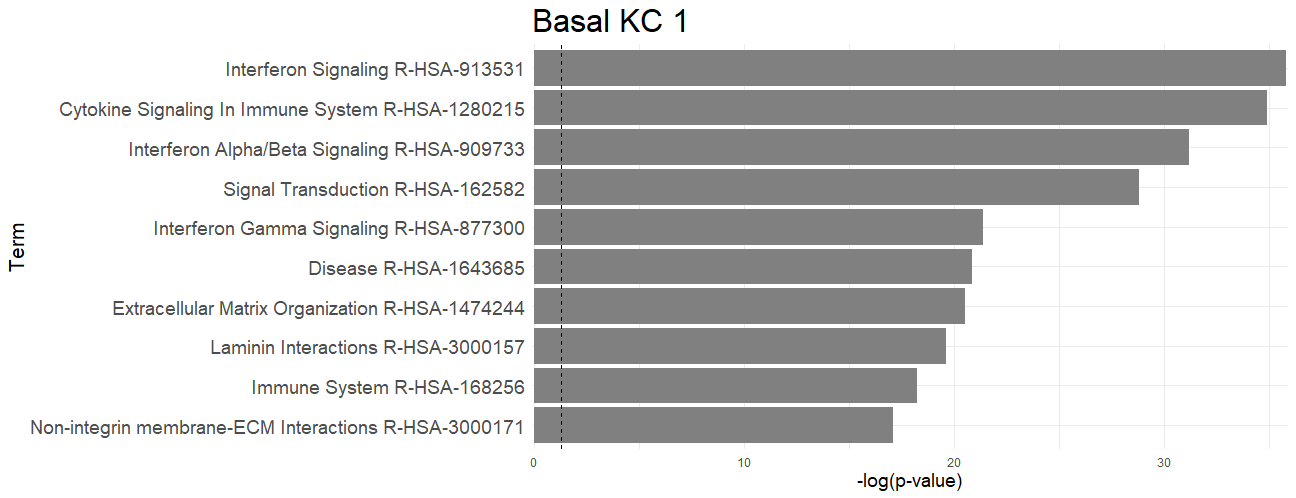

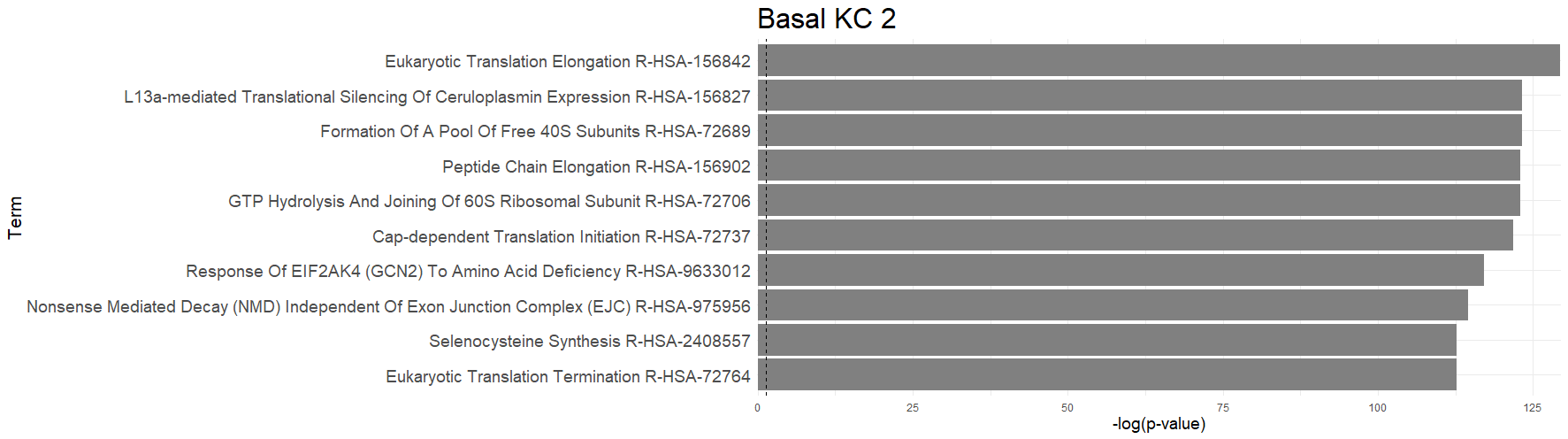

Basal KC 1

Basal KC 2

C

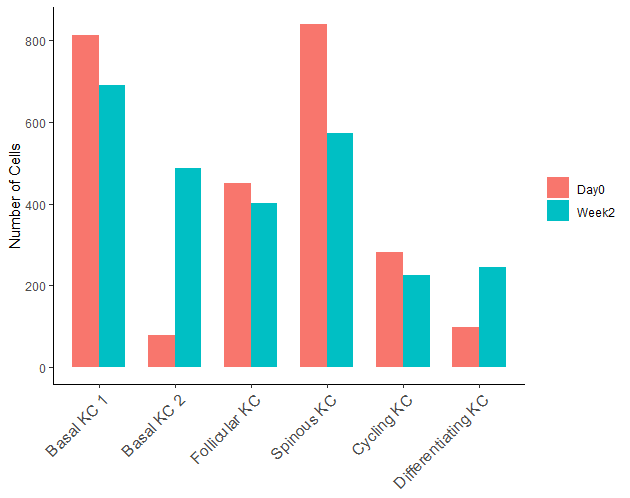

D

B

A

**Supplementary Figure 5.** (A) Top 5 keratinocyte markers for each keratinocyte subset identified in lesional LP skin. (B) Type II IFN and IFN-signature genes in different KC subsets. (C) Enriched Gene-Ontology pathways in “basal KC 1” and “basal KC 2” subsets compared to other keratinocytes. (D) Total cell numbers in each keratinocyte subsets in baseline (week 0, red) and week 2 (blue) (n=10 week 0 and n=10 week 2).
