## Supplementary Figure 6 for "Oral Baricitinib in the Treatment of Cutaneous Lichen Planus"

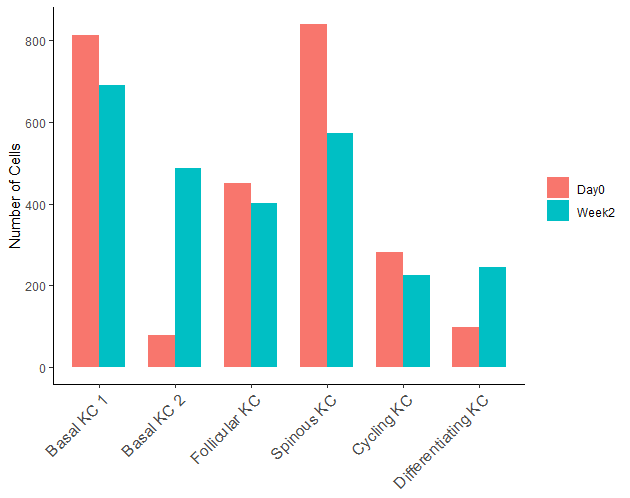

Week 0

Week 2

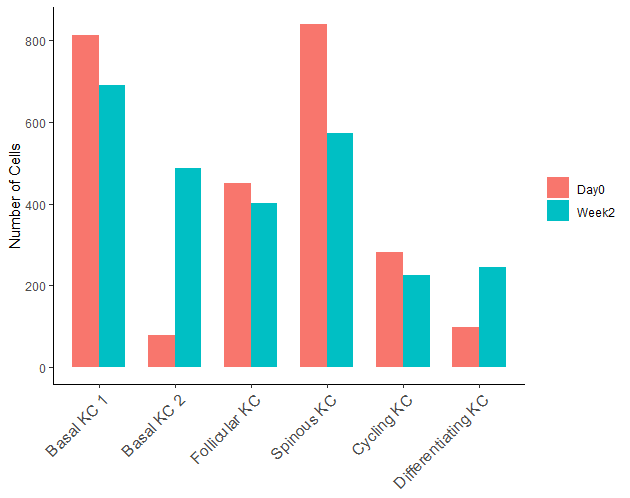
**Supplementary Figure 6.** (A) Marker genes for T cell subsets identified in lesional LP skin. (B) Spatial deconvolution of T-cell subsets in lesional LP samples (representative of n=9). (C) T cell numbers at baseline (red) and at week 2 after baricitinib treatment (n=10 for week 0 and week 2).

Week 0

Week 2

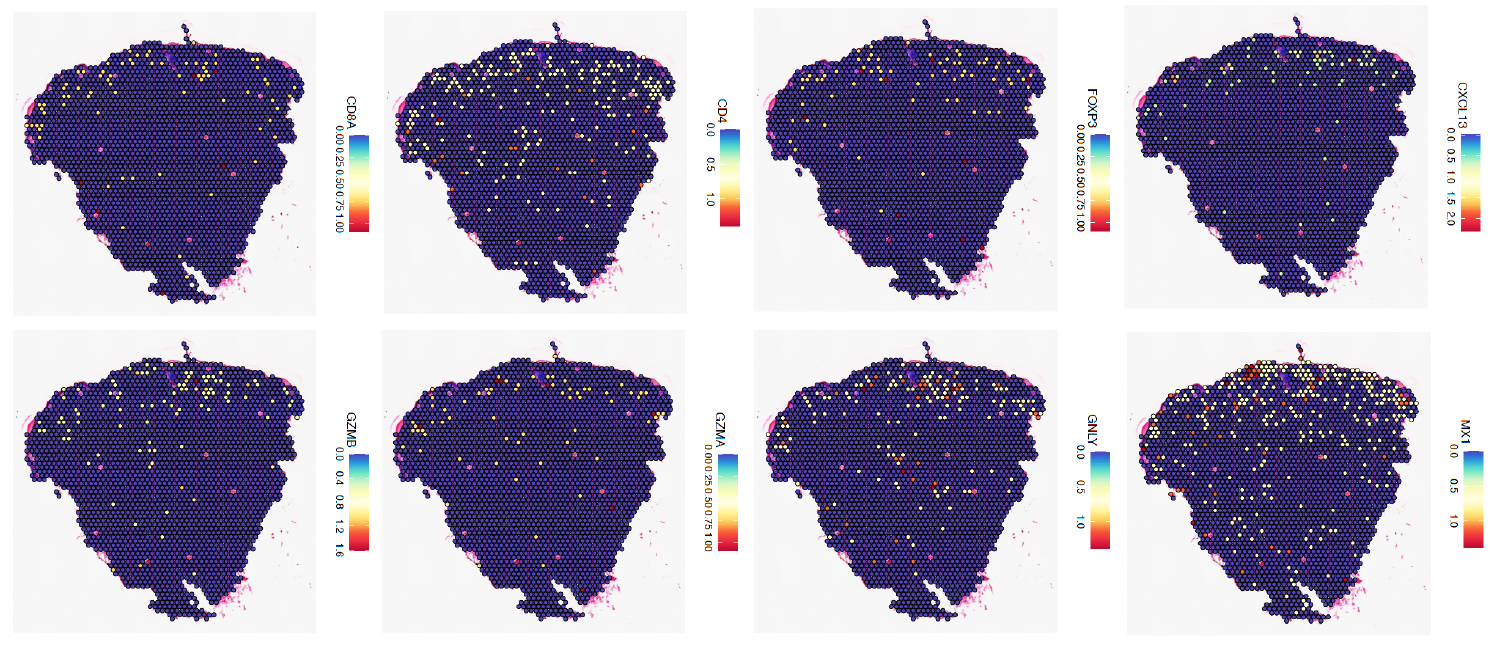

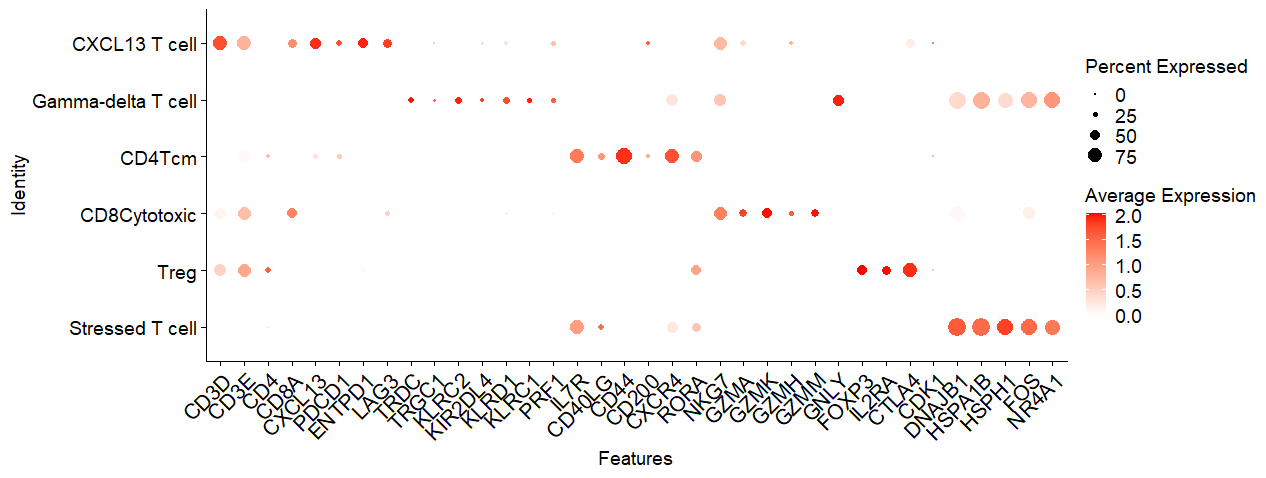

A

B

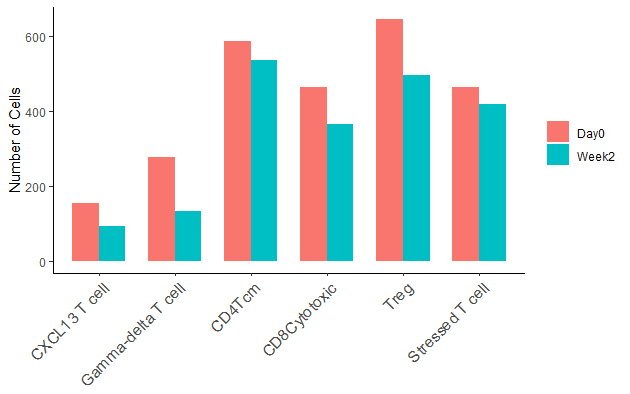

C
