## Supplementary Figure 8 for "Oral Baricitinib in the Treatment of Cutaneous Lichen Planus"

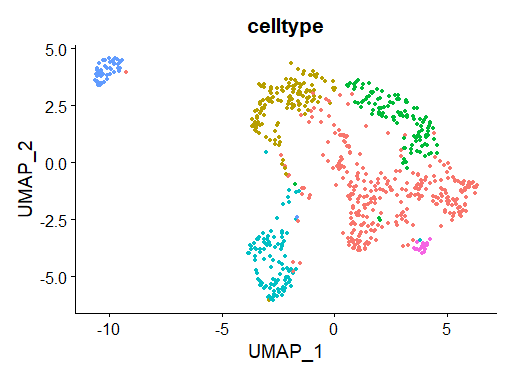

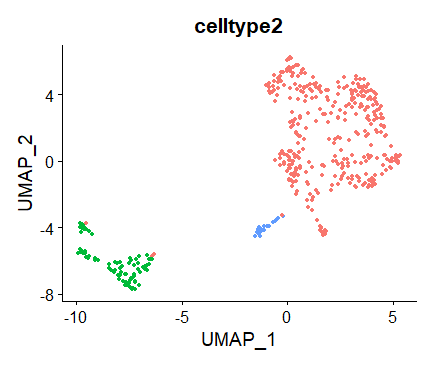

**SFRP2**

**FB**

**TNN**

**FB**

**SFRP4**

**FB**

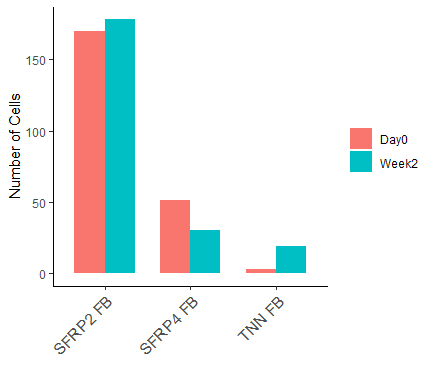

**Proliferating**

**Myeloid**

**M2-like Mac**

**CD1C DC**

**LAMP3 DC**

**CLEC9A DC**

**B Cell**

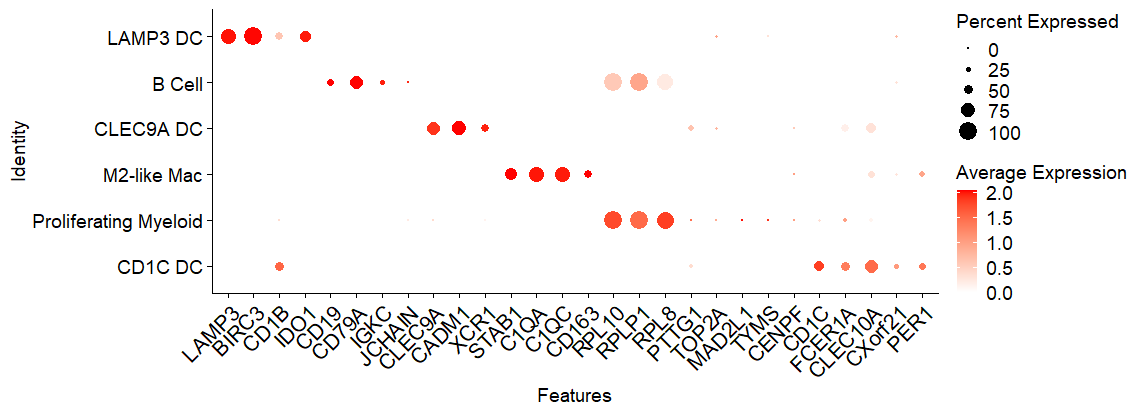

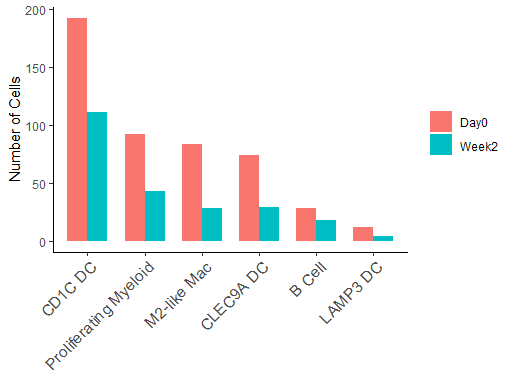

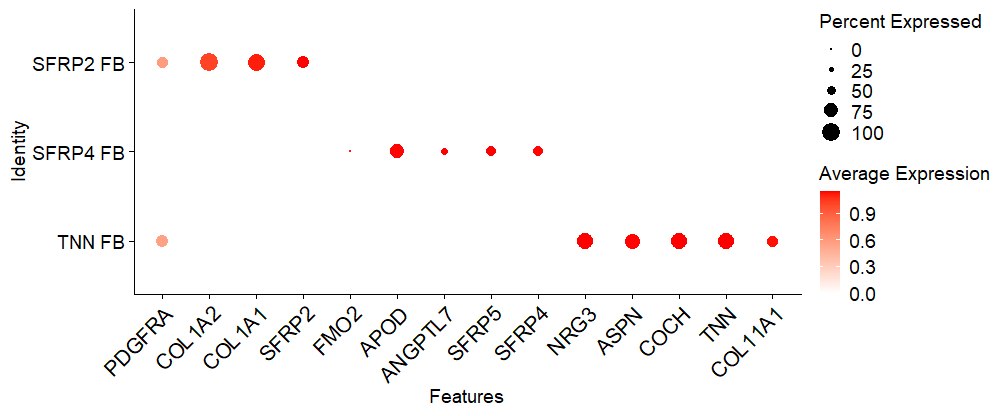

A

B

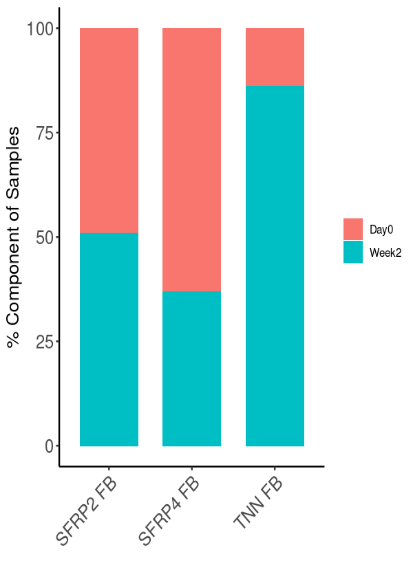

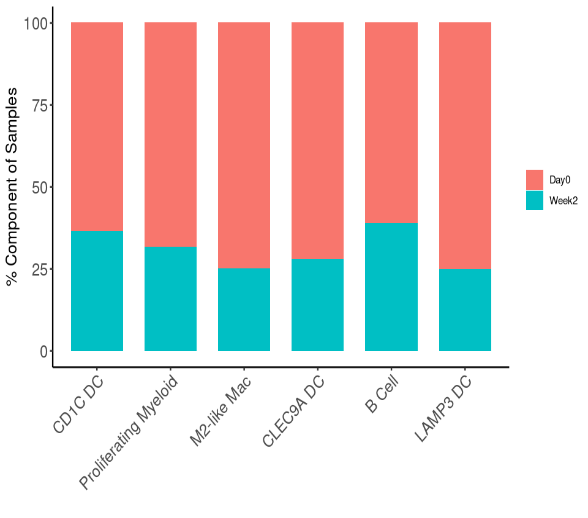

**Supplementary Figure 8.** (A) Myeloid and B cell populations in lesional LP skin. (B) Fibroblast subpopulations in LP skin. Data are shown for week 0 (Red) and week 2 (blue), along with the top 4 marker genes (n=10 at week 0 and n=10 at week 2).
