## Supplementary Figure 9 for "Oral Baricitinib in the Treatment of Cutaneous Lichen Planus"

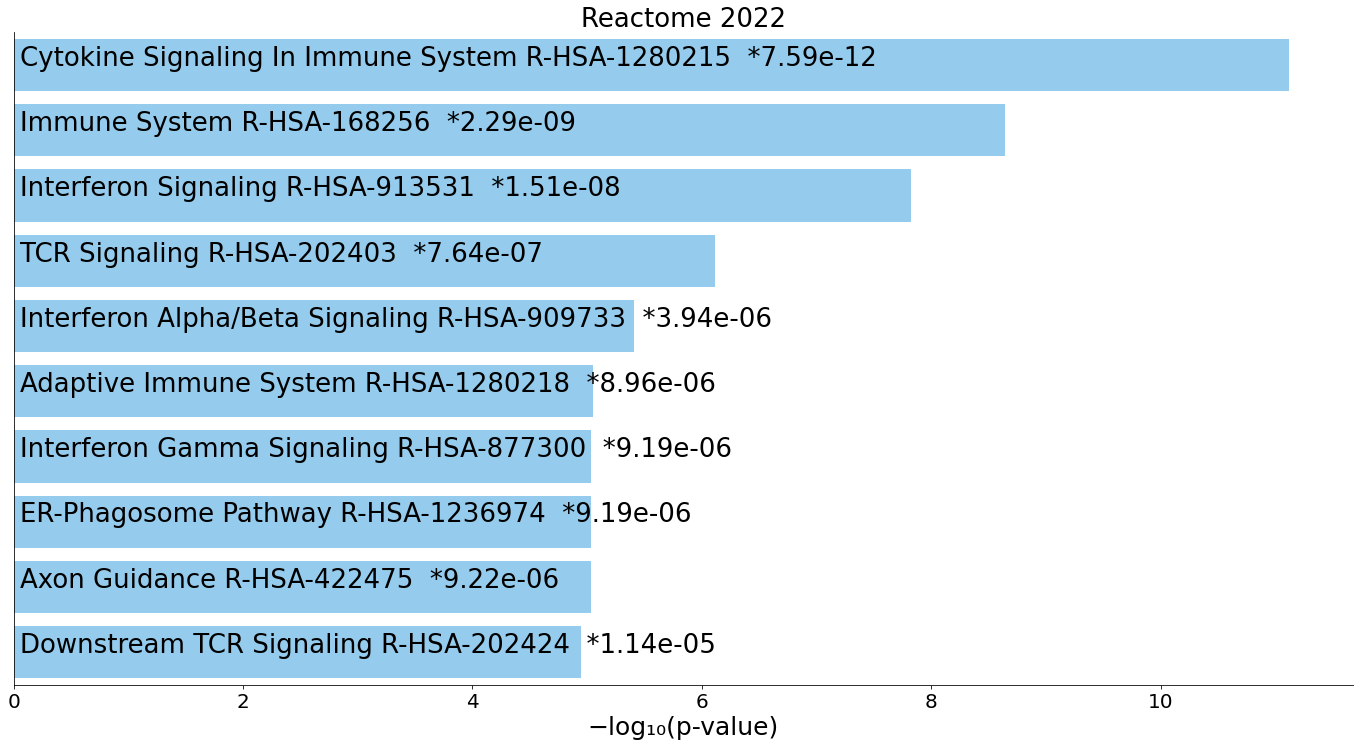

Genes decreased in myeloid cells week 2

Genes decreased in CD4 T cells week 2

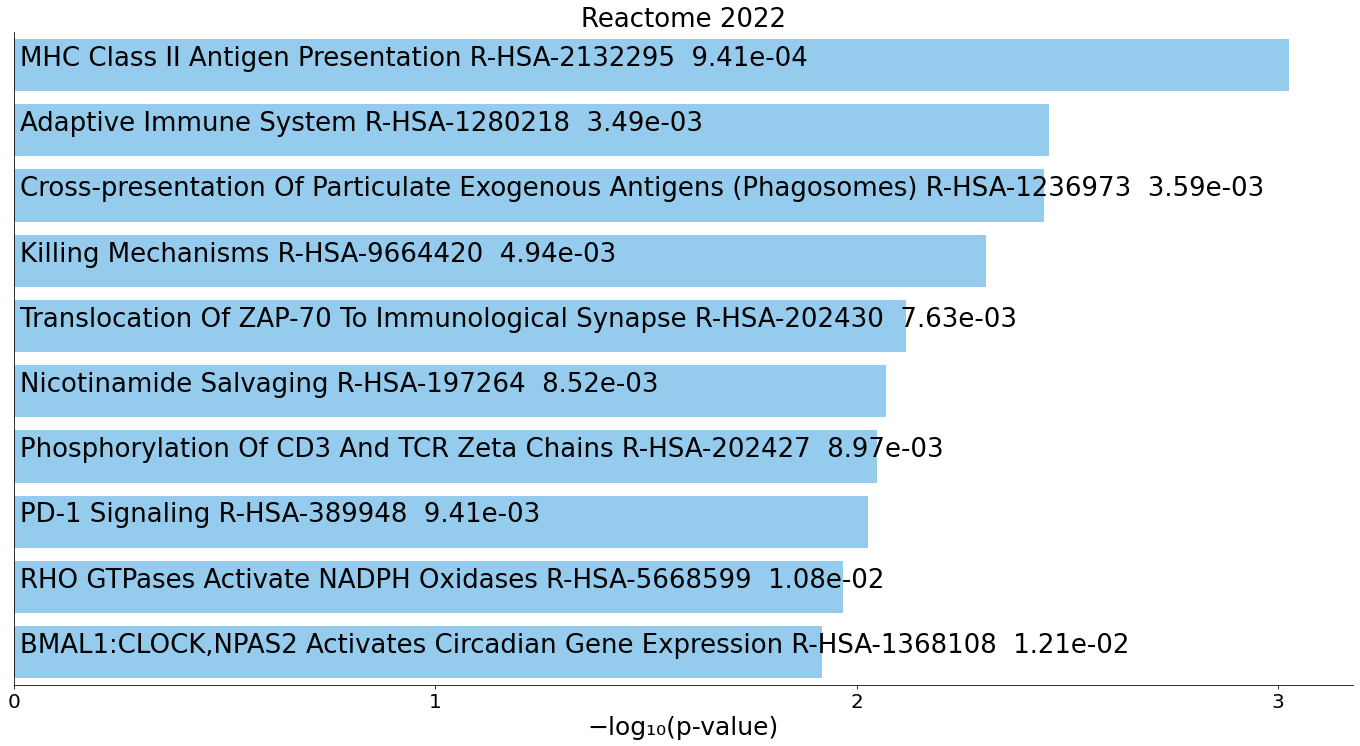

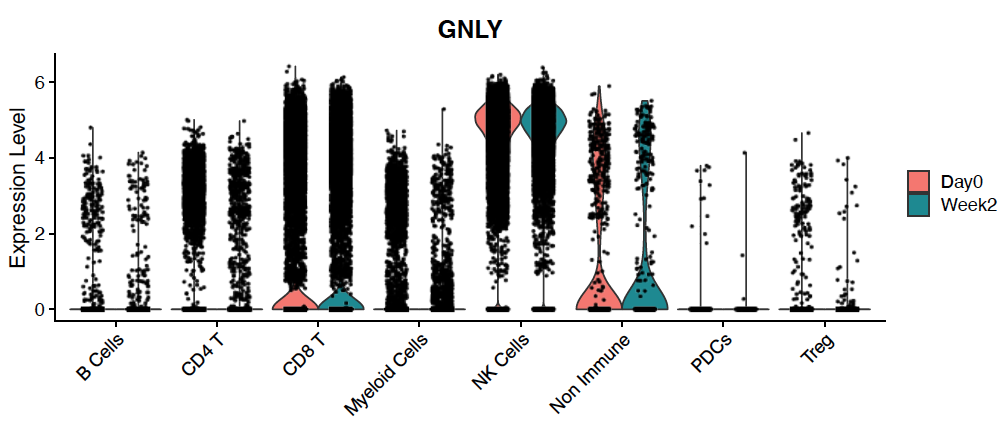

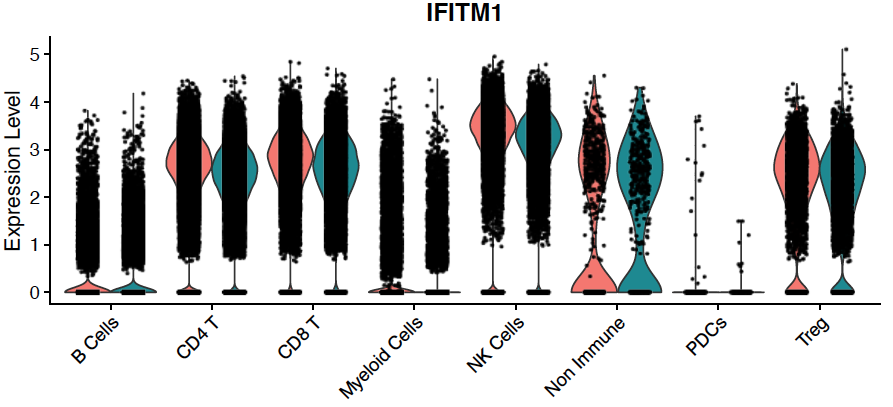

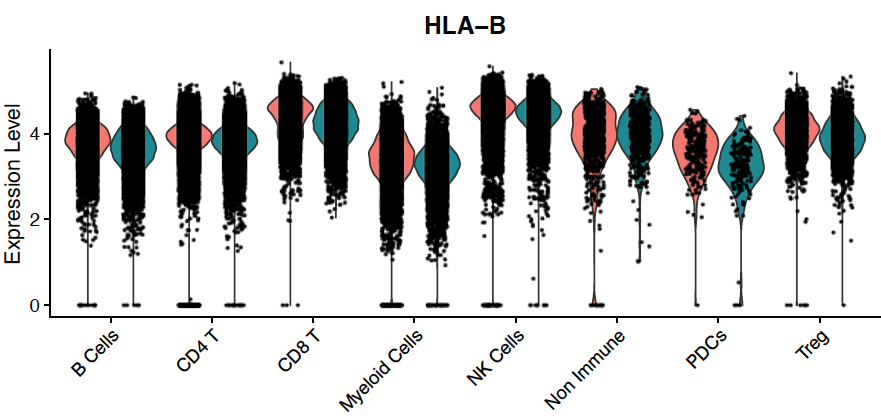

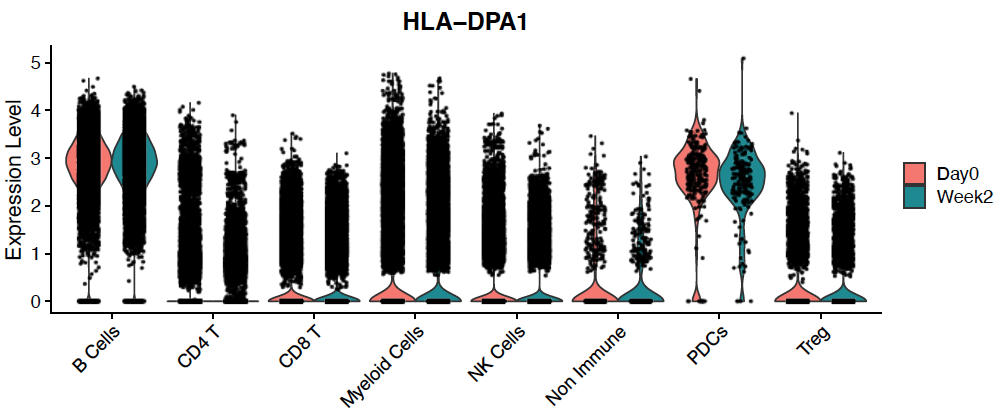

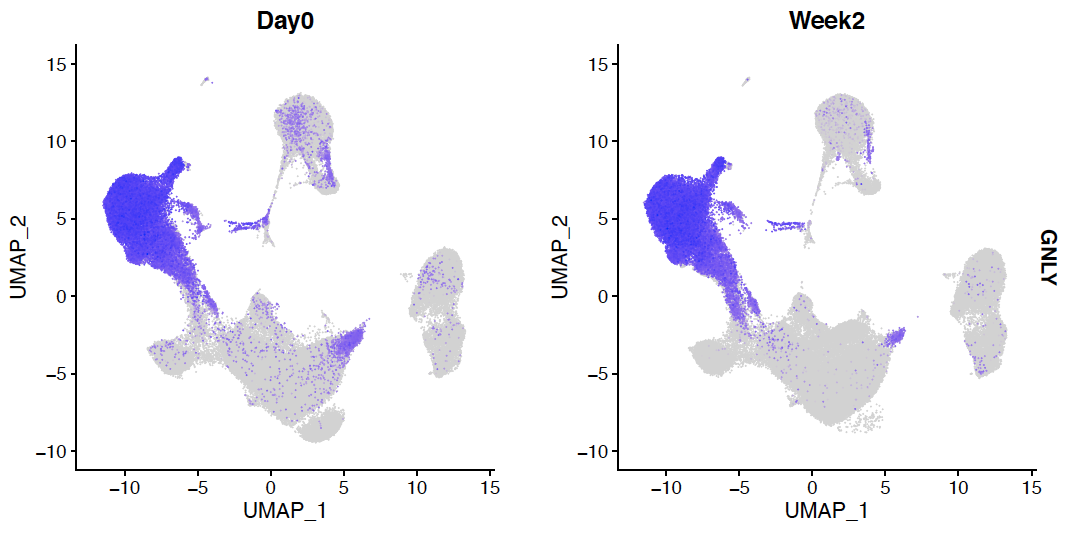

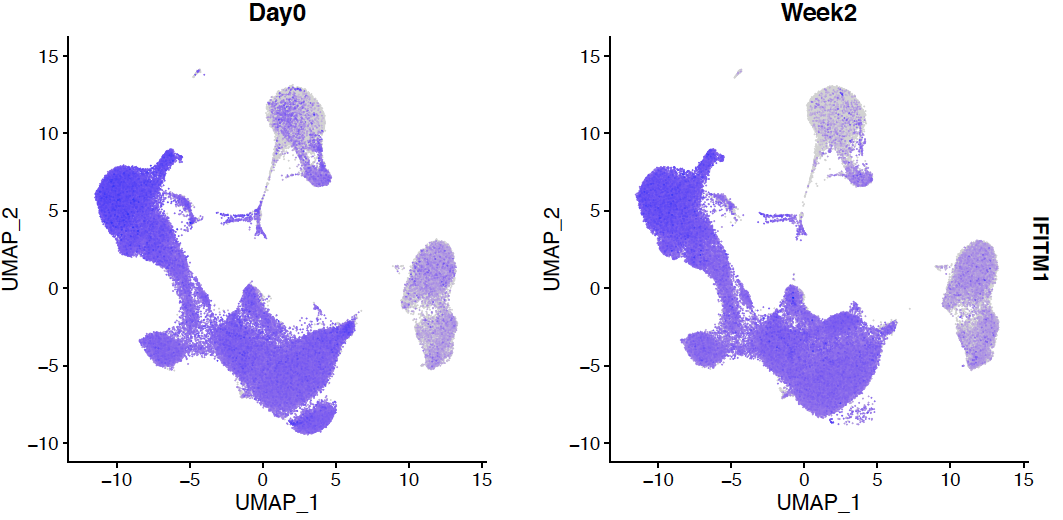

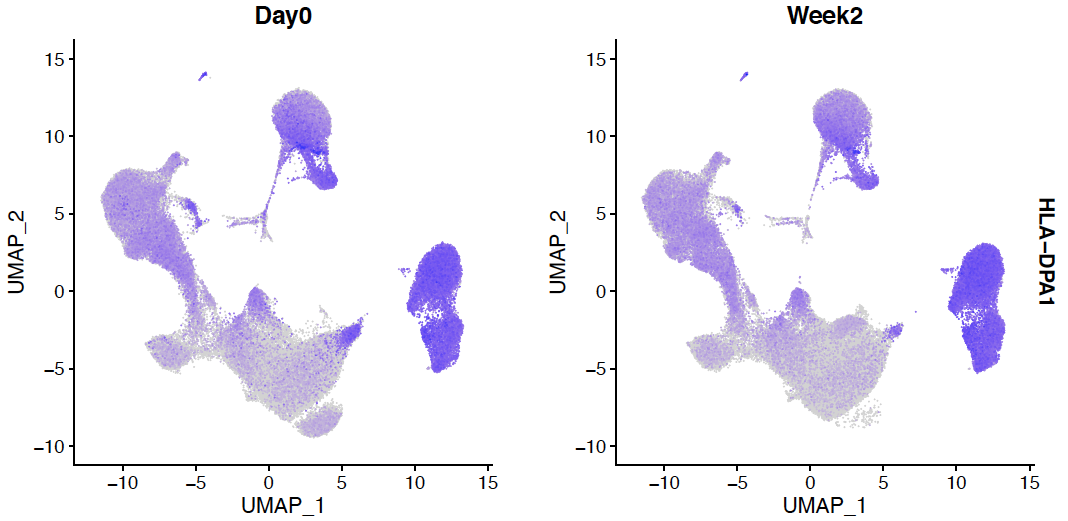

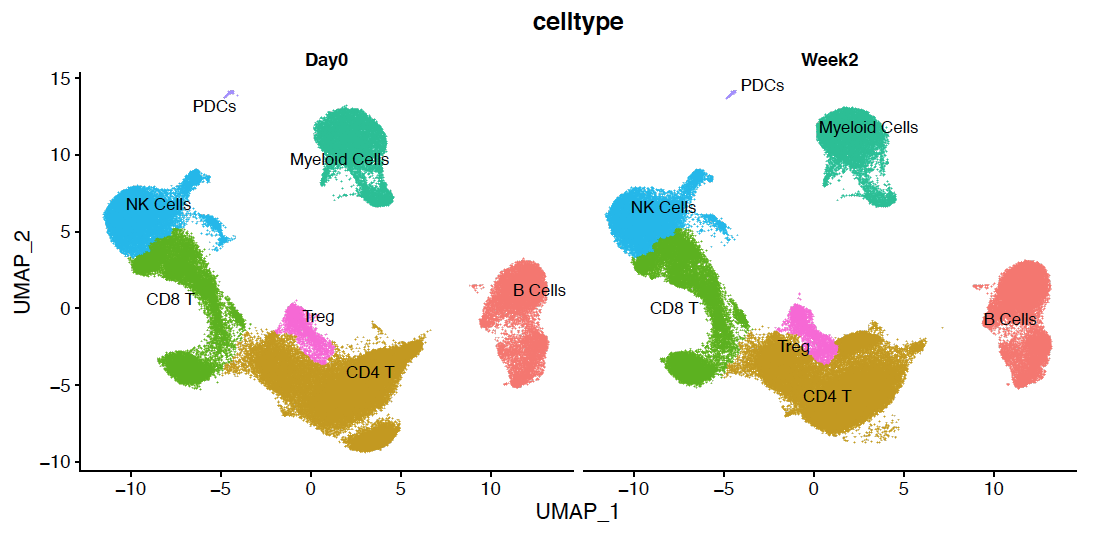

A

B

C

**Supplementary Figure 9.** (A) Changes in interferon signature genes (IFITM1), cytotoxic marker (GNLY), MHC class I (HLA-B), and MHC class II (DLA-DPA1) at baseline (day 0) vs. week 2 of treatment. (B) Single-cell sequencing of peripheral blood mononuclear cells (PBMCs) at baseline (day 0) and week 2. (C) Enriched gene ontology categories amongst DEGs in myeloid cells and CD4 T cells comparing week 2 vs baseline (n=10 at day 0, and n=10 at week 2).
