## Supplementary Table 1 for "Oral Baricitinib in the Treatment of Cutaneous Lichen Planus"

**Supplementary Table 1**. Physician Global Assessment (intention-to-treat analysis)

| **PGA** | **Week 1**  **(n=12)** | **Week 2**  **(n=12)** | **Week 3**  **(n=12)** | **Week 4**  **(n=12)** | **Week 8**  **(n=12)** | **Week 12**  **(n=12)** | **Week 16**  **(n=11)** | **Week 20**  **(n=11)** |
| --- | --- | --- | --- | --- | --- | --- | --- | --- |
| **Missing** | 4 | 0 | 4 | 0 | 0 | 1 | 0 | 0 |
| **Grade 0** | 0 (0%) | 0 (0%) | 0 (0%) | 0 (0%) | 6 (50.0%) | 5 (45.5%) | 5 (41.7%) | 4 (33..3%) |
| **Grade 1** | 0 (0%) | 1 (8.3%) | 2 (25.0%) | 4 (33.3%) | 1 (8.3%) | 4 (36.4%) | 5 (41.7%) | 5 (41.7%) |
| **Grade 2** | 0 (0%) | 3 (25.0%) | 2 (25.0%) | 3 (25.0%) | 1 (8.3%) | 0 (0%) | 0 (0%) | 1 (8.3%) |
| **Grade 3** | 3 (37.5%) | 5 (41.7%) | 3 (37.5%) | 2 (16.7%) | 2 (16.7%) | 2 (18.2%) | 0 (0%) | 1 (8.3%) |
| **Grade 4** | 3 (37.5%) | 2 (16.7%) | 1 (12.5%) | 3 (25.0%) | 2 (16.7%) | 0 (0%) | 1 (8.3%) | 0 (0%) |
| **Grade 5** | 2 (25.0%) | 1 (8.3%) | 0 (0%) | 0 (0%) | 0 (0%) | 0 (0%) | 0 (0%) | 0 (0%) |

Grade 0: Completely clear, no evidence of disease (100% improvement)

Grade 1: Almost clear, very significant clearance (>=90% to <100%)

Grade 2: Marked improvement, significant improvement (>=75% to <90%)

Grade 3: Moderate improvement, intermediate between slight and marked (>=50% to <75%)

Grade 4: Slight improvement, some improvement (>=25% to <50%) however, significant evidence of disease remains

Grade 5: No change, disease has not changed from baseline condition (+/- <25%)
