## Supplementary Table 2 for "Oral Baricitinib in the Treatment of Cutaneous Lichen Planus"

**Supplementary Table 2.** Primary and secondary endpoints at baseline and week 16 (per-protocol analysis)

|  | **Baseline** | **Week 16** | **Difference** | ***P* value** |
| --- | --- | --- | --- | --- |
| PGA |  |  |  |  |
| Responsive (n, %) |  | 90.9% |  |  |
| Non-responsive (n, %) |  | 9.1% |  |  |
| Total Body Lesion Count |  |  |  | 0.003 |
| Mean (SD) | 146.6 (168.9) | 8.0 (12.0) | -138.6 (164.0) |  |
| Range | 4.0 – 600.0 | 0.0 – 30.0 | -570.0 to -4.0 |  |
| mCAILS |  |  |  | 0.003 |
| Mean (SD) | 11.8 (2.7) | 1.1 (2.7) | -10.6 (3.1) |  |
| Range | 7.0 – 14.6 | 0.0 – 8.9 | -14.6 to -5.0 |  |
| BSA affected (%) |  |  |  | 0.003 |
| Mean (SD) | 4.2 (3.1) | 0.3 (0.9) | -3.9 (3.0) |  |
| Range | 0.1 – 10.0 | 0.0 – 3.0 | -9.8 to -0.1 |  |
| Pruritus NRS |  |  |  | 0.003 |
| Mean (SD) | 6.9 (2.4) | 1.1 (2.1) | -5.8 (2.1) |  |
| Range | 1.0 – 10.0 | 0.0 – 7.0 | -8.0 to -1.0 |  |
| Pruritus VAS |  |  |  | 0.003 |
| Mean (SD) | 6.6 (1.7) | 1.0 (2.0) | -5.6 (1.7) |  |
| Range | 2.7 – 8.8 | 0.0 – 6.3 | -7.7 to 2.5 |  |
| Pain NRS |  |  |  | 0.005 |
| Mean (SD) | 7.5 (1.6) | 1.2 (2.0) | -6.2 (2.1) |  |
| Range | 4.0 – 10.0 | 0.0 – 7.0 | -8.0 to 2.0 |  |
| Skindex-16 overall |  |  |  | 0.008 |
| Mean (SD) | 55.3 (19.4) | 9.0 (12.2) | -41.4 (13.6) |  |
| Range | 35.0 – 90.0 | 0.0 – 37.0 | -58.0 to -15.0 |  |
| Skindex-16 symptom |  |  |  | 0.005 |
| Mean (SD) | 15.6 (5.7) | 1.8 (3.0) | -13.6 (4.4) |  |
| Range | 4.0 – 24.0 | 0.0 – 10.0 | -19.0 to -4.0 |  |
| Skindex-16 emotional |  |  |  | 0.003 |
| Mean (SD) | 28.8 (8.6) | 6.0 (7.0) | -22.8 (7.0) |  |
| Range | 17.0 – 42.0 | 0.0 – 17.0 | -34.0 to 10.0 |  |
| Skindex-16 functional |  |  |  | 0.012 |
| Mean (SD) | 9.9 (8.8) | 2.0 (3.4) | -6.0 (5.6) |  |
| Range | 0.0 – 29.0 | 0.0 – 10.0 | -15.0 to 0.0 |  |
