## Supplementary Table 3 for "Oral Baricitinib in the Treatment of Cutaneous Lichen Planus"

**Supplementary Table 3**. Primary and secondary endpoints in dose escalation cohort

|  | **Week 16** | **Week 28** | **Difference** | ***P* value** |
| --- | --- | --- | --- | --- |
| PGA |  |  |  |  |
| Clear; PGA 0 (n, %) | 0, 0.0% | 3, 60.0% |  |  |
| Almost clear; PGA 1 (n, %) | 4, 80.0% | 1, 20.0% |  |  |
| Total Body Lesion Count |  |  |  | 0.138 |
| Mean (SD) | 17.6 (12.2) | 5.0 (10.6) | - 12.6 (17.6) |  |
| Range | 2.0 – 30.0 | 0.0 – 24.0 | -30.0 to 11.0 |  |
| mCAILS |  |  |  | 0.593 |
| Mean (SD) | 2.4 (3.7) | 1.4 (2.6) | -1.1 (4.3) |  |
| Range | 0.0 – 8.9 | 0.0 – 6.0 | -8.0 to 3.9 |  |
| BSA affected (%) |  |  |  | 0.343 |
| Mean (SD) | 0.7 (1.3) | 0.3 (0.4) | -0.4 (1.3) |  |
| Range | 0.1 – 3.0 | 0.0 – 1.0 | -2.5 to 0.9 |  |
| Pruritus NRS |  |  |  | 0.357 |
| Mean (SD) | 2.2 (2.8) | 1.2 (1.3) | -1.0 (2.2) |  |
| Range | 0.0 – 7.0 | 0.0 – 3.0 | -4.0 to 2.0 |  |
| Pruritus VAS |  |  |  | 0.225 |
| Mean (SD) | 2.0 (2.7) | 0.8 (1.5) | -1.2 (1.6) |  |
| Range | 0.0 – 6.3 | 0.0 – 3.5 | -3.0 to 0.3 |  |
| Pain NRS |  |  |  | 0.197 |
| Mean (SD) | 2.2 (2.8) | 1.2 (1.6) | -1.0 (1.6) |  |
| Range | 0.0 – 7.0 | 0.0 – 4.0 | -3.0 to 1.0 |  |
| Skindex-16 overall |  |  |  | 1.000 |
| Mean (SD) | 12.8 (16.4) | 16.8 (19.9) | 4.0 (12.3) |  |
| Range | 1.0 – 37.0 | 0.0 – 41.0 | -5.0 to 18.0 |  |
| Skindex-16 symptomatic |  |  |  | 0.891 |
| Mean (SD) | 3.6 (3.8) | 3.6 (4.4) | 0.0 (2.0) |  |
| Range | 1.0 – 10.0 | 0.0 – 11.0 | -2.0 to 3.0 |  |
| Skindex-16 emotional |  |  |  | 0.285 |
| Mean (SD) | 7.4 (6.9) | 11.8 (17.0) | 6.8 (12.4) |  |
| Range | 0.0 – 17.0 | 0.0 – 36.0 | -4.0 to 24.0 |  |
| Skindex-16 functional |  |  |  | 0.655 |
| Mean (SD) | 3.0 (4.8) | 3.6 (4.2) | 0.8 (5.4) |  |
| Range | 0.0 – 10.0 | 0.0 – 10.0 | -5.0 to 8.0 |  |
