## Supplementary Table 4 for "Oral Baricitinib in the Treatment of Cutaneous Lichen Planus"

**Supplementary Table 4**. Physician Global Assessment in dose escalation cohort

| **PGA** | **Week 16**  **(n=5)** | **Week 20**  **(n=5)** | **Week 24**  **(n=5)** | **Week 28**  **(n=5)** | **Week 32**  **(n=5)** |
| --- | --- | --- | --- | --- | --- |
| **Grade 0** | 0 (0%) | 1 (20.0%) | 1 (20.0%) | 3 (60.0%) | 1 (20.0%) |
| **Grade 1** | 4 (80.0%) | 3 (60.0%) | 2 (40.0%) | 1 (20.0%) | 1 (20.0%) |
| **Grade 2** | 0 (0%) | 0 (0%) | 0 (0%) | 0 (0%) | 0 (0%) |
| **Grade 3** | 0 (0%) | 1 (20.0%) | 1 (20.0%) | 0 (0%) | 2 (40.0%) |
| **Grade 4** | 1 (20.0%) | 0 (0%) | 0 (0%) | 1 (20.0%) | 0 (0%) |
| **Grade 5** | 0 (0%) | 0 (0%) | 1 (20.0%) | 0 (0%) | 1 (20.0%) |
