## Supplementary Table 5 for "Oral Baricitinib in the Treatment of Cutaneous Lichen Planus"

**Supplementary Table 5.** Adverse Events

|  | **Possible/Probable/Definite** | **Unrelated/Unlikely** | **Total** |
| --- | --- | --- | --- |
| **Adverse Event** |  |  |  |
| Ankle cramp | 0 (0.0%) | 1 (9.1%) | 1 (8.3%) |
| Blood cell count decreased | 1 (100.0%) | 0 (0.0%) | 1 (8.3%) |
| Chest tenderness | 0 (0.0%) | 1 (9.1%) | 1 (8.3%) |
| COVID | 0 (0.0%) | 1 (9.1%) | 1 (8.3%) |
| Lichen planus flare | 0 (0.0%) | 1 (9.1%) | 1 (8.3%) |
| Migraine with pain above right eye to the cheek | 0 (0.0%) | 1 (9.1%) | 1 (8.3%) |
| Night sweats | 0 (0.0%) | 1 (9.1%) | 1 (8.3%) |
| Pain lower right leg and right side | 0 (0.0%) | 1 (9.1%) | 1 (8.3%) |
| Pain upper right side | 0 (0.0%) | 1 (9.1%) | 1 (8.3%) |
| Shortness of breath, chest pain | 0 (0.0%) | 1 (9.1%) | 1 (8.3%) |
| Upper respiratory tract infection/nasopharyngitis (common cold) | 0 (0.0%) | 1 (9.1%) | 1 (8.3%) |
| Visual field changes, floaters and intermittent flashing left eye | 0 (0.0%) | 1 (9.1%) | 1 (8.3%) |
| **Serious Adverse Event** |  |  |  |
| Missing | 0 | 1 | 1 |
| No | 1 (100.0%) | 10 (100.0%) | 11 (100.0%) |
| Yes | 0 (0.0%) | 0 (0.0%) | 0 (0.0%) |
| **Severity Grade** |  |  |  |
| Mild | 1 (100.0%) | 6 (54.5%) | 7 (58.3%) |
| Moderate | 0 (0.0%) | 3 (27.3%) | 3 (25.0%) |
| Severe | 0 (0.0%) | 2 (18.2%) | 2 (16.7%) |
